# Mpox vaccine acceptance and uptake in Africa: a systematic review and meta-analysis (1970-2024)

**DOI:** 10.1101/2025.10.12.25337821

**Authors:** Fabrice Zobel Lekeumo Cheuyem, Andreas Ateke Njoh, Chabeja Achangwa, Otfried Kistner, Rick Tchamani, Jessy Goupeyou-Youmsi, Davy Roméo Takpangdo-Legrand, Sory Kourouma, Mazou Ngou Temgoua

## Abstract

**Background:** Mpox remains a significant public health threat in Africa, with recent outbreaks driven by newly emergent clades I and II. Vaccination is a critical intervention for outbreak control, yet evidence on vaccine acceptance and uptake across the continent has not been comprehensively synthesized. This study aimed to determine the pooled prevalence and determinants of mpox vaccine acceptance and uptake in Africa.

**Methods:** We conducted a systematic review and meta-analysis following PRISMA guidelines, registered with PROSPERO (CRD420251126033). We searched PubMed, Scopus, Web of Science, CINAHL, ScienceDirect, and African Journals Online from 1970 to August 2025. Data were extracted using a standardized form, and quality was assessed with Joanna Briggs Institute tools. Pooled estimates were calculated using random-effects generalized linear mixed models with Probit-Logit transformation. Subgroup analyses and meta-regressions explored heterogeneity by region, population, setting, and time period.

**Results:** Of 9,748 records screened, 35 studies were included. The overall mpox vaccine acceptance rate was 53.55% (95% CI: 46.16–60.79), with high heterogeneity (I^2^=98%). Central Africa showed moderate acceptance at 54.17% (95% CI: 20.82–84.16), Eastern Africa at 54.16% (95% CI: 42.43– 65.44), while Western Africa was lower at 50.11% (95% CI: 39.94–60.27). Acceptance was highest in Southern Africa (67.43%; 95% CI: 61.85–72.67). Healthcare workers’ acceptance was 51.63% (95% CI: 39.37–63.70) and medical students’ was 46.17% (95% CI: 38.53–54.01), both lower than the general population (62.46%; 95% CI: 52.25–71.66). Actual vaccine uptake was 20.94% (95% CI: 10.06–38.56), varying significantly by country, with the Democratic Republic of the Congo at 20.01% (95% CI: 7.45–43.75). A significant decline occurred after 2022 (pre-2022: 36.0% [95% CI: 19.74–56.26]; post-2022: 3.4% [95% CI: 0.56–17.96]). Key determinants of acceptance included higher mpox knowledge, trust in health authorities, prior vaccination history, and free vaccine access.

**Conclusion:** There is a substantial gap between mpox vaccine acceptance and actual uptake in Africa, with a significant decline in coverage since 2022. Strategies to enhance vaccination must address both demand and supply challenges, including equitable distribution and integration into routine immunization systems. These findings underscore the need for context-specific, multi-level interventions to translate willingness into actual vaccine uptake.

## Background

Mpox (monkeypox) is an infectious viral disease caused by the monkeypox virus, a double-stranded DNA virus belonging to the *Orthopoxvirus* genus within the *Poxviridae* family. This family also includes the viruses responsible for smallpox (*variola*), cowpox, and vaccinia [1].. The infection can cause a painful rash, enlarged lymph nodes, and fever [2].

The mpox virus is causing concurrent outbreaks in several African countries [3]. The Democratic Republic of Congo (DRC) bears the highest global burden, with travel-related cases that can potentially lead to global outbreaks [4,5]. In August 2024, the Africa CDC declared mpox a Public Health Emergency of Continental Security, while the World Health Organization (WHO) previously declared it a Public Health Emergency of International Concern in July 2023. These declarations followed a global outbreak driven by newly emerged, virulent clades I (Ia, Ib) and II (IIa, IIb) of the mpox virus [6,7]. By June 2025, several African nations had reported recent community transmission of the clade Ib mpox virus [8].

To effectively combat the mpox pandemic, global health authorities have recommended that countries adopt and implement preventive strategies. These strategies include avoiding close skin-to-skin contact with people who have an mpox-like rash, avoiding close contact with animals that could carry the virus, frequently washing hands with soap or an alcohol-based sanitizer, and vaccination [9– 11]. Vaccination is considered one of the most effective interventions for preventing and controlling the unprecedented spread of mpox, which has led to a case fatality rate of over 4% in some Sub-Saharan African countries, such as the DRC [4,12].

To prevent mpox, vaccination is recommended both as a primary preventive measure for high-risk individuals and as post-exposure prophylaxis for known contacts. Following the initial outbreak, global containment efforts have prioritized vaccination to curb the virus’s spread [13]. The JYNNEOS vaccine, a replication-deficient vaccinia virus vaccine, is currently being used for this purpose. It was approved by the Food and Drug Administration (FDA) after being proven to be the safest available substitute for the previously used ACAM2000 vaccine.

The ACAM2000 vaccine was linked to dangerous side effects, including myopericarditis and even death [14,15]. In contrast, the most frequently reported side effects of the JYNNEOS vaccine were milder and easier to manage, typically including injection site reactions, headache, myalgia, chills, and nausea [16].

Beyond the fear of potential adverse effects, vaccine hesitancy which is recognized as one of the top ten global health threats [17–20], is fueled by misinformation, attitudes and perceptions such as the fear of needles and doubts about a vaccine’s efficacy and usefulness [21,22]. Additionally, studies in Africa have identified several sociodemographic characteristics associated with vaccines acceptance and uptake[19,20,23–26].

In this regard, the need to develop and implement an mpox vaccination toolkit has become crucial. Such a toolkit offers a set of practical guidance, training, and information resources to help relevant stakeholders in African countries prepare, plan, implement, and monitor mpox vaccination programs [27]. Moreover, community engagement is essential for the successful implementation of these programs. By fostering trust, ensuring cultural sensitivity, tailoring interventions, and building sustainable health systems, it is a key component of a successful vaccination strategy [28]. Key strategies for engagement include involving trusted local actors and community representatives in decision-making, disseminating accessible information through diverse platforms, and actively collecting social and behavioral insights to adapt strategies and combat misinformation [29].

However, the primary challenge to controlling mpox in Africa is the limited access to vaccines, which is hindered by both supply and distribution issues [30]. Another significant challenge is identifying high-risk adults for vaccination, particularly due to the sexual transmissibility of Clade I mpox strains. A third major hurdle is designing and implementing vaccination programs that effectively reach at-risk children under five, who face the highest mortality risk. This challenge is compounded by the need to ensure that their parents are willing to accept vaccination for their children [31].

Several studies conducted in Africa provide insight into the use of the mpox vaccine to combat recurrent outbreaks. These studies indicate varying levels of vaccine acceptance and uptake [24,32–41,41–48]. A key knowledge gap exists regarding the pooled acceptance and coverage rates for the mpox vaccine, as well as the socio-behavioral factors influencing them. This uncertainty hinders the development of tailored, subregional or country-specific vaccination strategies. To address these limitations, we conducted a systematic review and meta-analysis to estimate the prevalence and synthesize the determinants of mpox vaccine acceptance and uptake across Africa, stratifying by subregion, country, and key populations, including the general public and healthcare workers.

## Methods

### Study design

This review was conducted in accordance with the Preferred Reporting Items for Systematic Reviews and Meta-Analyses (PRISMA) guidelines and was registered with the International Prospective Register of Systematic Reviews (PROSPERO ID: CRD420251126033)[49,50].

### Eligibility criteria

This systematic review and meta-analysis included all published studies from Africa that reported on mpox vaccination uptake and acceptance based on condition, context, and population [51]. Accordingly, we included original, full-text articles that reported on any of our study’s outcomes of interest: rates of intention to vaccinate against mpox, rates of vaccine uptake, or factors associated with mpox vaccine acceptance or uptake.

We excluded studies that were only available as abstracts or focused solely on conditional acceptance, such as willingness to pay for the vaccine. We also did not include studies that failed to report our primary outcomes, measured outcomes only with continuous variables without providing prevalence rates, or were strictly clinical trials of the mpox vaccine.

### Article searching strategy

We identified published research through a systematic search of online databases, including PubMed, Scopus, CINAHL, Web of Science, ScienceDirect, and African Journals Online (AJOL). The search process involved screening titles and abstracts using a combination of keywords and Medical Subject Headings (MeSH). We used Boolean operators like “AND” and “OR” to refine our search with terms such as:(Mpox OR Monkeypox) AND (vaccine OR vaccination OR acceptance) AND (African countries). To ensure comprehensive coverage, we also conducted a manual search on Google Scholar for publications not indexed in the primary databases and screened the reference lists of all included studies for relevant articles. The final search was completed on August 07, 2025.

### Data extraction

We used Zotero software (version 7.0.16) to retrieve and de-duplicate references from our literature search. One investigator (FZLC) then used a pre-designed Microsoft Excel 2016 form to extract key study characteristics. The form captured the first author’s name, study year, country, study design, participant type, setting, sampling technique, the number of individuals willing to receive the vaccine, the number who received it, the total sample size and the factors associated with vaccine acceptability and/or uptake. The collected data was then cross-checked by two other investigators (RT and AAN).

### Critical appraisal (quality assessment) of included studies

Two authors independently assessed the relevance and quality of each article (FZLC and AC). Any disagreements between the reviewers were resolved through discussion with a third author (MNT) to reach a consensus. The Joanna Briggs Institute (JBI) quality assessment tool was used to evaluate the quality of studies included [52]. For cross-sectional studies, assessment criteria encompassed studies clear definition of inclusion criteria, comprehensive descriptions of study subjects and settings, validity and reliability of exposure measurements, use of objective, standardized criteria for outcome assessment, identification potential confounding factors, implementation of appropriate strategies to address them, and the appropriateness of statistical methods. For cohort study, this evaluation examined group comparability between exposed and unexposed populations, consistency and accuracy of exposure measurement, identification and adjustment for confounding factors, and confirmation of outcome-free status at baseline. Each criterion was scored as 1 (yes) or 0 (no or unclear). The overall risk of bias was categorized as low (>50%), moderate (>25-50%), or high (≤25%).

### Study outcome

The primary outcomes of this systematic review and meta-analysis were mpox vaccine acceptance and mpox vaccine uptake. Vaccine acceptance was defined as the unconditional willingness to receive a free mpox vaccine if offered [13,24]. Vaccine uptake referred to individuals who had either self-reported receiving the vaccine, presented a vaccination card, or showed a characteristic smallpox vaccine scar [32].

### Statistical analysis and synthesis

The rates for vaccine acceptance and uptake were calculated by dividing the number of people who met each criterion by the total number of participants and multiplying by 100.

To explore the variability in results, subgroup analyses were conducted based on several factors: study period (before and after the 2022 global outbreak), country, setting, participant type, sampling technique, and sample size (categorized as above or below the median value). Countries were grouped by WHO African region [53], including the Western (Algeria, Ghana, Nigeria), Eastern (Ethiopia, Kenya, Uganda), Southern (South Africa), and Central (Cameroon, Central African Republic, DRC) regions. The analysis also included a non-WHO Afro region (Northern Africa), which comprised Morocco and Egypt. Univariable and multivariable meta-regression models were then used to determine if these characteristics explained the variation in the pooled estimates.

The *I*^2^ statistics was used to assess heterogeneity between studies, with categories defined as low (<25%), moderate (25–75%), or high (>75%). A random-effects model was chosen to pool the estimates, anticipating significant heterogeneity. The Generalized Linear Mixed Models (GLMM), coupled with the Probit-Logit Transformation (PLOGIT), were utilized for their effectiveness in handling meta-analyses of binary data and accommodating studies with 0% or 100% event rates without requiring continuity corrections [54]. Statistical significance was set at a p-value of <0.05. All analyses were conducted using the ‘meta’ package in R Statistics version 4.4.2 [55]. Chart were generated online using MapChart online application [56].

### Publication bias and sensitivity test

To evaluate potential publication bias among the included studies, we used funnel plots. The symmetry of the inverted funnel shape suggested the absence of publication bias the Egger’s and Begg’s tests [57,58]. A p-value□>□0.05 indicating no statistically significant evidence of publication bias. Sensitivity analysis was conducted by iteratively excluding one study at a time to assess the robustness of the findings.

## Results

A total of 9748 entries were obtained through an online database search and a manual search. After removing 749 duplications, 8999 distinct entries underwent title and abstract review, followed by a complete text evaluation for suitability. Ultimately, 35 research papers satisfied the inclusion standards and were incorporated into the meta-analysis (Fig. 1).

**Fig. 1:**
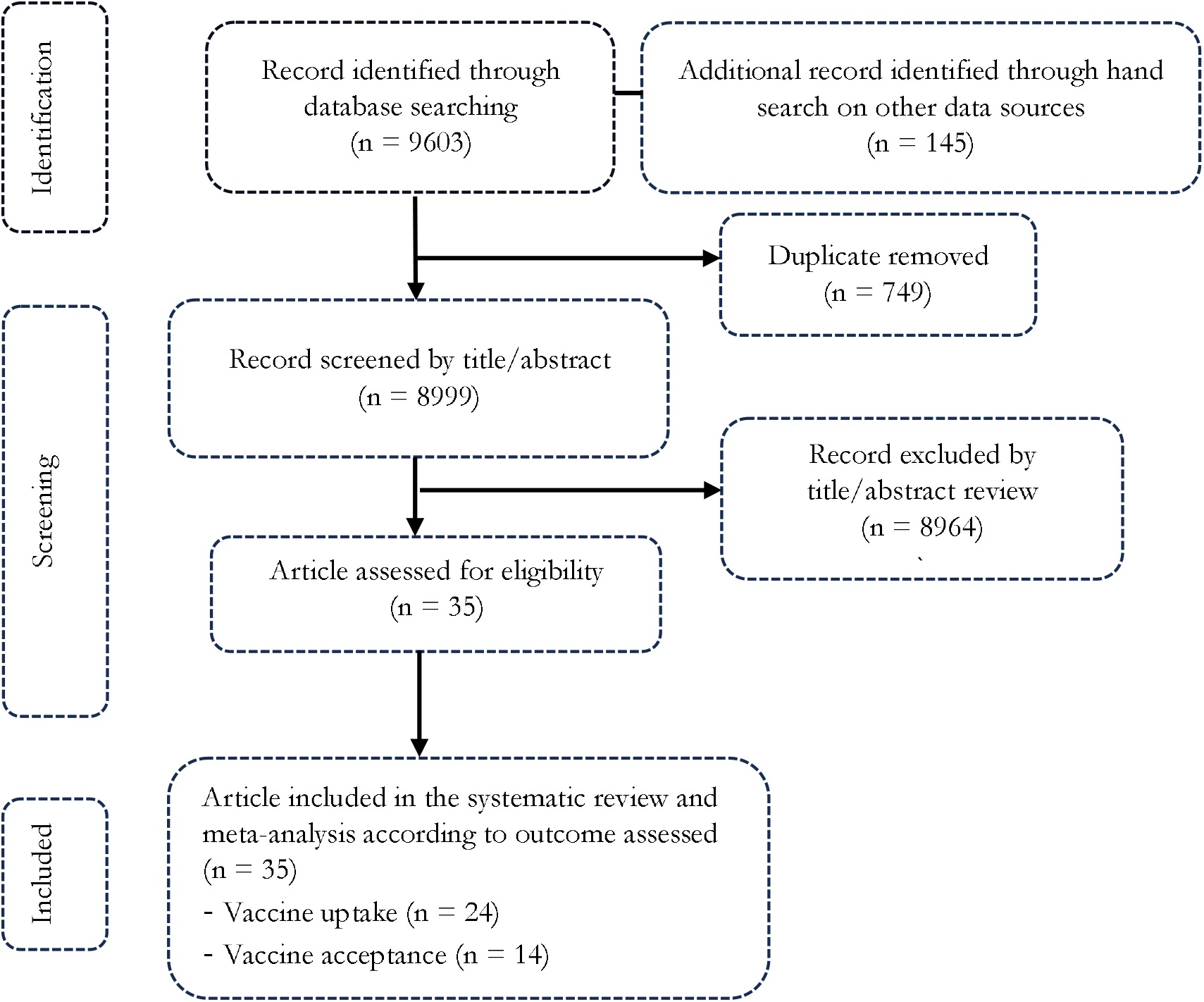
PRISMA diagram flow of studies included in the meta-analysis

### Studies selection

### Characteristics of studies included

This analysis of mpox vaccination studies across Africa reveals several key patterns: the evidence base was dominated by cross-sectional studies (100% of included studies) [16,24–26,32–48,59–72], primarily focused on the DRC (45% of studies) [24,32–41,41–48], with most (84%) using non-probabilistic sampling methods)[16,24,32–48,60–67,69,71]. The research shows vaccine acceptance (VA) [16,24–26,41,42,63,64,66,67,67–70,72] and uptake (VU) [26,32–48,59–62,65,69,70,72] outcomes or both [26,41,42,69,70,72], with studies interested healthcare workers (21%) [16,25,41,42,62,63,66,68,70,72] and online data collection (16%) in recent years [16,25,63,64,67,69,71]. Most studies were rated low risk of bias (87%) [16,24–26,32–45,47,48,59– 61,65–72] (Additional Files 1, Supplementary Table 1).

### Mpox vaccine acceptance

This meta-analysis reveals moderate overall mpox vaccine acceptance in Africa of 53.55% (95% CI: 46.16-60.79), though with high heterogeneity (*I*^2^=98%; *p* ⍰ 0.001) indicating significant variation between studies. The highest prevalence was observed in study conducted in DRC (85%; 95%CI: 57.19-98.22) [42] and lowest in Ethiopia (28.84%; 95%CI: 25.62-32.23) [25] (Fig. 2).

**Fig. 2:**
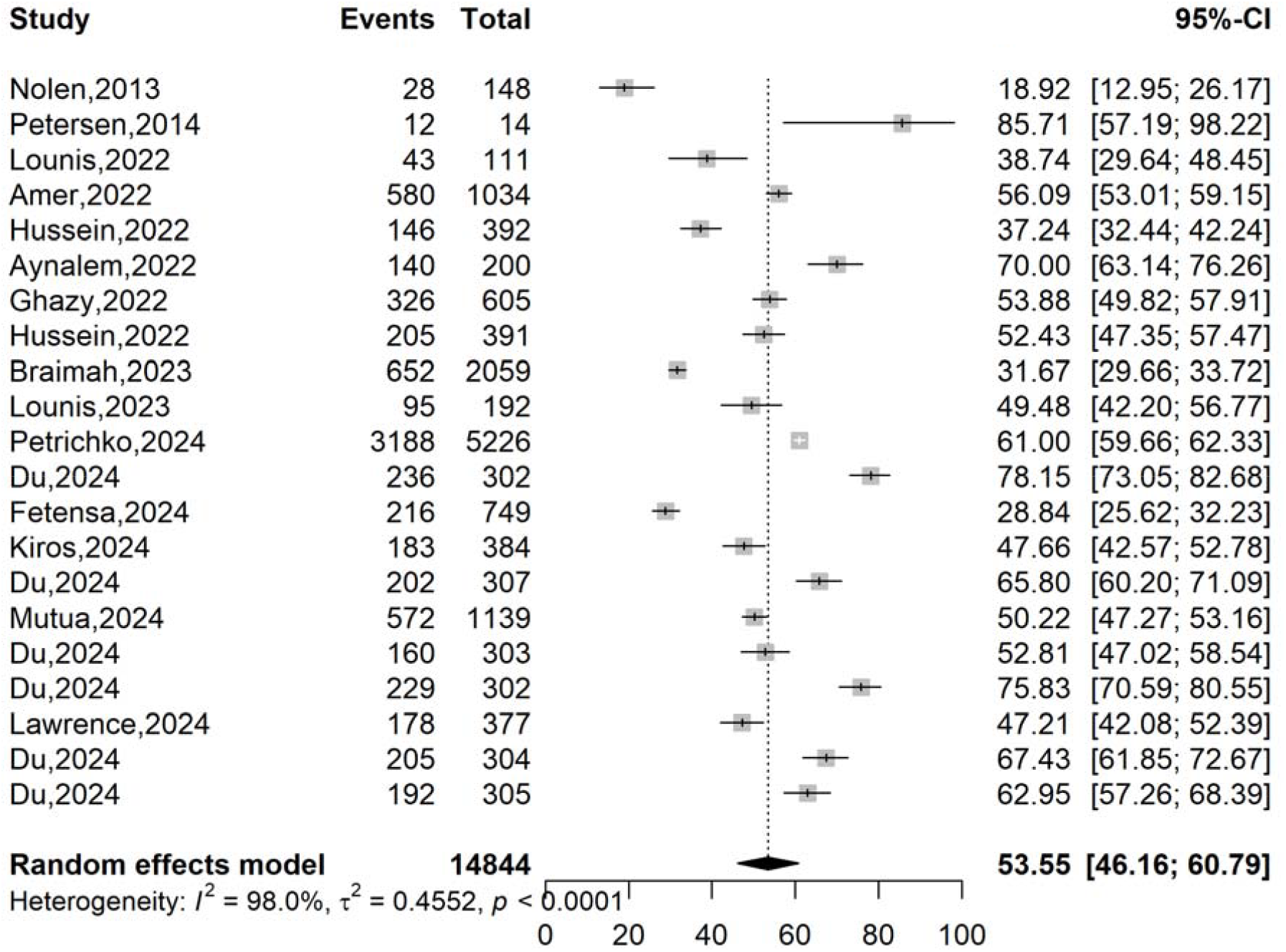
Pooled estimates of mpox vaccine acceptance rate in Africa, 2013-2024

While some countries like South Africa (67.43%; n = 1), Uganda (62.95%; n = 1) [69] showed relatively highest acceptance, others such as and Ghana (42.27%; n = 2) [16,26], Algeria (44.98%; n = 2 studies) [63,64] and Ethiopia (48.53%; n = 3)[25,68,70] demonstrated lower rates. There was a significant difference between subgroup pooled estimates between countries (*p* = 0.001) and WHO Afro regions (*p* = 0.021) (Fig. 3; Supplementary Fig. 2 and 5).

**Fig. 3:**
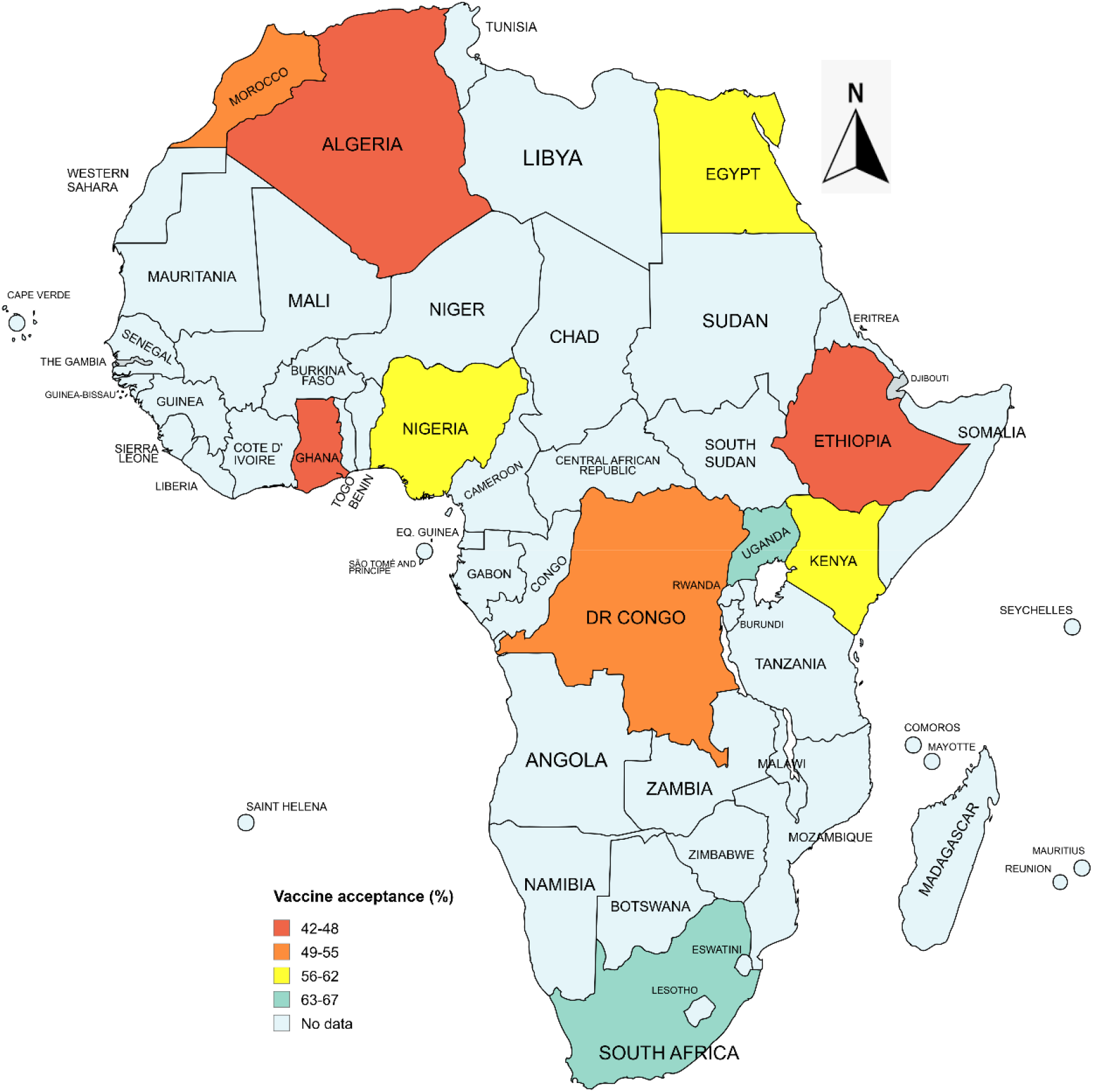
Map of the pooled vaccine acceptance rate by country, Africa, 1970-2024 (See Additional File 2, Supplementary Fig. 2)

### Subgroup analysis of vaccine acceptance rate

Subgroup assessment revealed a suboptimal mpox vaccination acceptance was observed across Africa (31.68–67.43%), with high heterogeneity (*I*^2^>89.7%, all *p* < 0.001) in all of subgroups and indicating substantial variation between studies. While acceptance increased slightly post-2022 (54.35% vs. 52.11%), populations combining general public and healthcare workers presented the lowest rate (34.68%; n = 2 studies). Hospital based studies (55.21) highlighted higher acceptance compare to studies in communities (47.32%; n = 4 studies). Notably, healthcare workers (51.63%; n = 7 studies) and medical students (46.17%; n = 3 studies) showed low vaccination rates (Table 1, Additional Files 1, Supplementary Fig. 1-8)

**Table 1:** Subgroup analysis of mpox vaccination acceptance rate in Africa, 2013-2024.

| Subgroup | Sample size | Coverage (%) | 95% CI limits |  | Number of studies | Heterogeneity statistic<sup>1</sup> |  |
| --- | --- | --- | --- | --- | --- | --- | --- |
|  |  |  | Lower | Upper |  | I<sup>2</sup>(%) | p-value |
| **Study period<sup>2</sup>** |  |  |  |  |  |  |  |
| < 2022 | 162 | 52.11 | 9.33 | 92.00 | 2 | 94 | < 0.001 |
| 2022 + | 14,682 | 54.35 | 47.79 | 60.76 | 19 | 98 | < 0.001 |
| **WHO Afro region** |  |  |  |  |  |  |  |
| Yes | 12,813 | 52.76 | 44.45 | 60.91 | 17 | 98.2 | < 0.001 |
| No | 2,031 | 56.80 | 41.25 | 71.12 | 4 | 97.2 | < 0.001 |
| **WHO Afro zone<sup>3</sup>** |  |  |  |  |  |  |  |
| Central | 5,388 | 54.17 | 20.82 | 84.16 | 3 | 97.6 | < 0.001 |
| Western | 4,037 | 50.11 | 39.94 | 60.27 | 7 | 97.8 | < 0.001 |
| Eastern | 3,084 | 54.16 | 42.43 | 65.44 | 6 | 97.6 | < 0.001 |
| Southern | 304 | 67.43 | 61.85 | 72.67 | 1 | - | - |
| Northern/non-WHO Afro region | 2,031 | 56.80 | 41.25 | 71.12 | 4 | 97.2 | < 0.001 |
| **Setting** |  |  |  |  |  |  |  |
| Community | 7,447 | 47.32 | 22.41 | 73.64 | 4 | 99.5 | < 0.001 |
| Hospital | 1,995 | 55.21 | 46.15 | 63.94 | 4 | 91.4 | < 0.001 |
| Online | 5,402 | 55.35 | 47.01 | 63.39 | 13 | 97 | < 0.001 |
| **Participant** |  |  |  |  |  |  |  |
| General population | 9,108 | 62.46 | 52.25 | 71.66 | 8 | 98.9 | < 0.001 |
| General population and Healthcare worker | 753 | 34.68 | 14.70 | 62.05 | 2 | 98 | < 0.001 |
| Healthcare worker | 2,869 | 51.63 | 39.37 | 63.70 | 7 | 96.6 | < 0.001 |
| Medical student | 975 | 46.17 | 38.53 | 54.01 | 3 | 89.7 | < 0.001 |
| Teacher | 1,139 | 50.22 | 47.27 | 53.16 | 1 | - | - |
| **Sampling** |  |  |  |  |  |  |  |
| Non-probabilistic | 11,075 | 56.59 | 48.16 | 64.28 | 16 | 95.4 | < 0.001 |
| Probabilistic | 3,769 | 44.60 | 31.94 | 58.01 | 4 | 97.5 | < 0.001 |
| **Sample size** |  |  |  |  |  |  |  |
| □ 500 | 4,032 | 56.51 | 47.28 | 65.32 | 15 | 95.5 | < 0.001 |
| 500 + | 10,812 | 46.63 | 36.68 | 56.86 | 6 | 99.2 | < 0.001 |

<sup>1</sup> Random effects model; CI: Confidence Interval; <sup>2</sup> Period before and after the mpox global outbreak.; <sup>3</sup> Western: Algeria, Ghana, and Nigeria; Eastern: Ethiopia, Kenya, and Uganda; Southern: South Africa; Central: Democratic Republic of Congo

### Meta-regression analysis of vaccine acceptance rate

The meta-regression assessing potential source of heterogeneity did not identify any significant factors influence the heterogeneity of vaccine acceptance estimate between studies (Table 2).

**Table 2:**
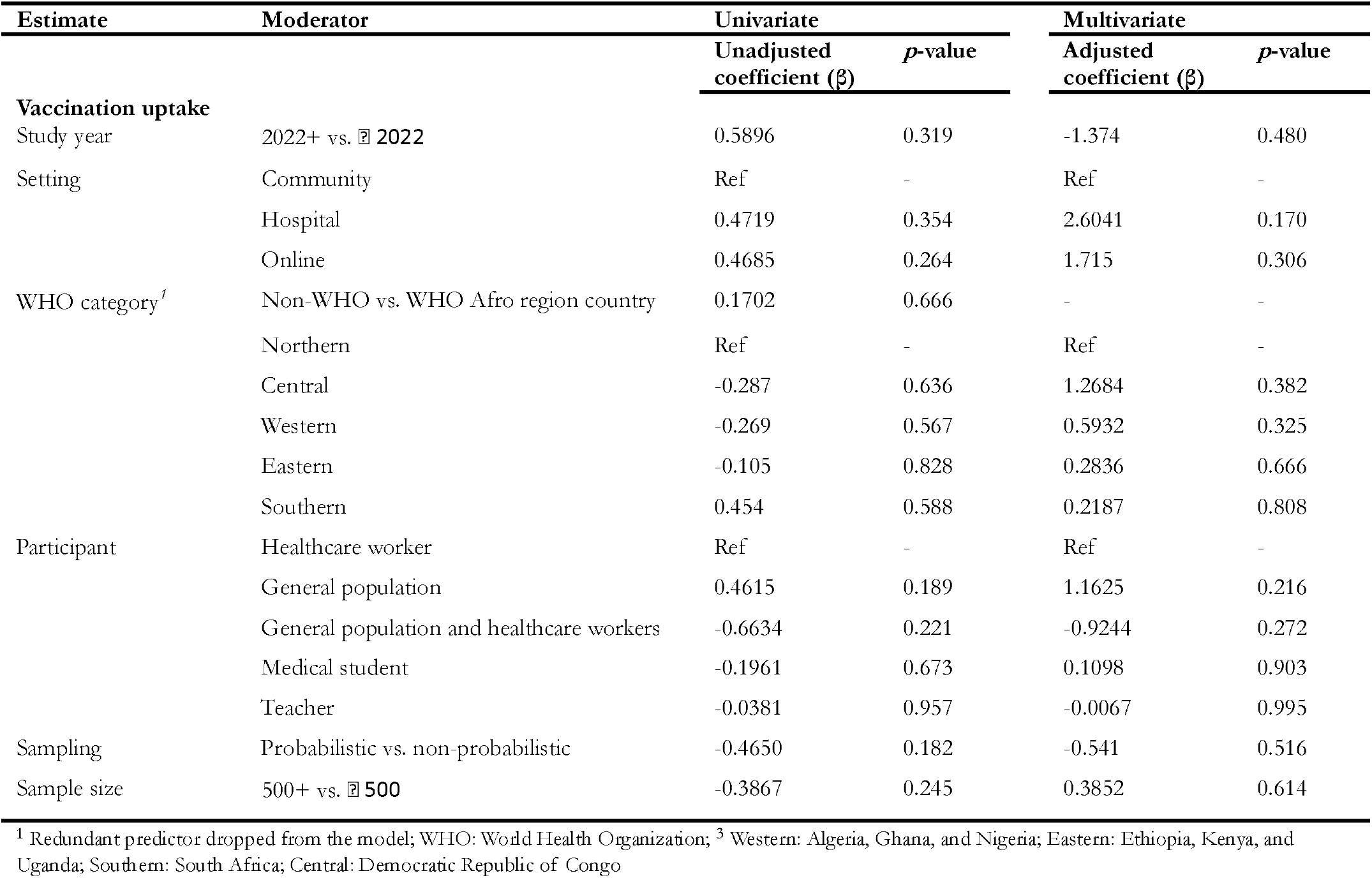
Meta-regression analysis of mpox vaccination coverage and acceptance in DRC, 1970-2024.

### Mpox vaccine uptake

The pooled vaccination coverage in Africa was 20.94% (95% CI: 10.06-38.56). The meta-analysis reveals extreme variability across studies, with coverage rates ranging from 0% (Kiros *et al*., 2024, Vakaniaki *et al*., 2024) [48,70] to 94.08% (Ladnyi 1970) [32], demonstrating substantial heterogeneity (*I*^*2*^ = 99.7%, *p* <0.001). While some historical studies showed high coverage such as Jezek *et al*., 1984 (91.07%), and Mande *et al*., 2019: 92.96%), more recent data indicate concerning declines (Du *et al*., 2024: 5.76%, Brosius *et al*., 2024: 2.27%) (Fig. 4).

**Fig. 4:**
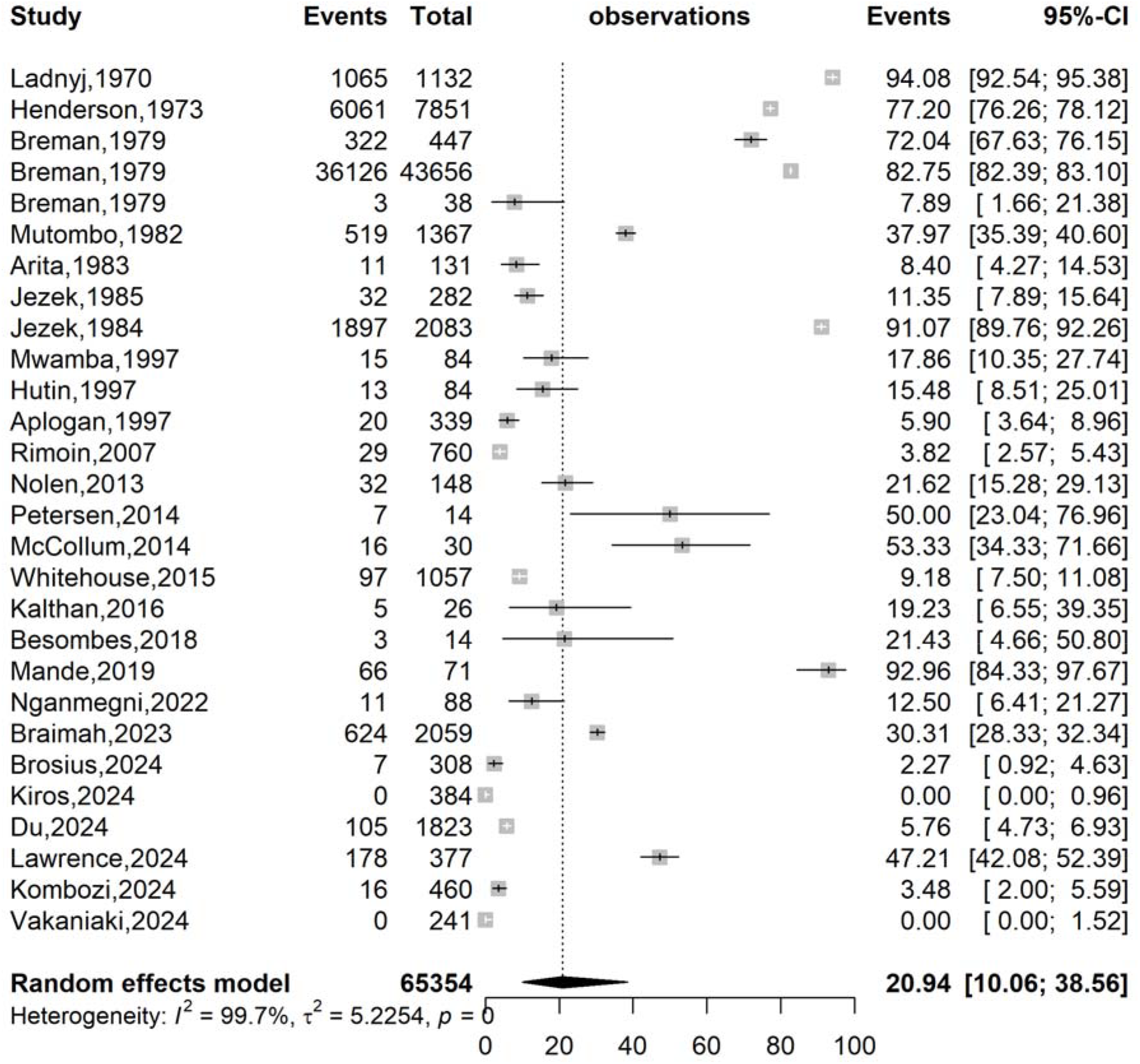
Pooled mpox vaccination coverage estimates in Africa, 1970-2024

**Fig. 5:**
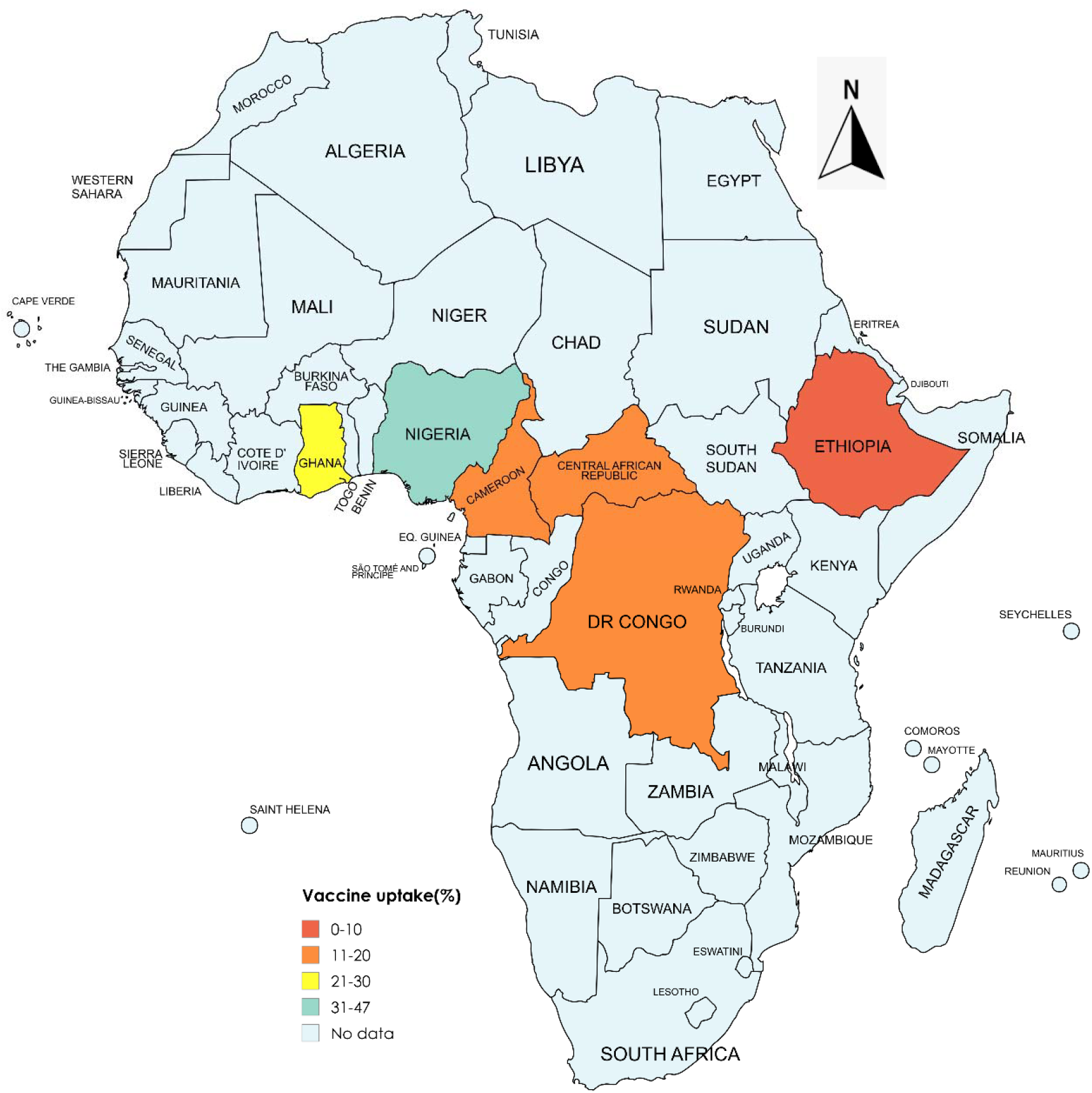
Map of the vaccine coverage trends by country, Africa, 1970-2024 (See Additional file 3, Supplementary Fig. 2)

This subgroup analysis of mpox vaccination uptake in Africa reveals disparities across countries. The DRC shows particularly inconsistent results (pooled 20.01%, 95% CI:7.45-43.75), ranging from 0% coverage (Vakaniaki 2024) to 94.08% (Ladnyj1970), suggesting significant temporal differences [32,48]. Multi-country studies demonstrate moderate but highly variable coverage (42.89%; CI:12.42-79.91), while low uptake appears in recent data from Ethiopia (0%, n = 1 study), Cameroon (12.5%; n = 1 study), and Central African Republic (20%; n = 2 studies). The significant subgroup differences (*p* < 0.001) highlight that geographic location influence vaccination rates (Fig. 5; Additional File 3, Supplementary Fig. 2).

This subgroup analysis of mpox vaccination uptake rate reveals a significant (p <0.001) decline from pre-2022 (36.0%; n = 20 studies) to post-2022 periods (3.4%; n = 8 studies) with the lowest rates observed in Eastern Africa (0%; n = 1 study), hospital-based studies (1.70%; n = 3), among healthcare workers (5.5%; n = 3 studies), and demonstrating high heterogeneity within most subgroups (*I*^2^>97.5%, *p* <0.001). Meanwhile, multi-regions studies (42.89%; n = 5 studies) and western Africa showed higher coverage (38.1%; n = 2 studies) (Table 3; Additional Supplementary Fig. 1-7).

**Table 3:** Subgroup analysis of mpox vaccination uptake rate in Africa, 2013-2024.

| Subgroup | Sample size | Coverage (%) | 95% CI limits |  | Number of studies | Heterogeneity statistic<sup>1</sup> |  |
| --- | --- | --- | --- | --- | --- | --- | --- |
|  |  |  | Lower | Upper |  | *I*<sup>2</sup>(%) | *p*-value |
| **Study period<sup>2</sup>** |  |  |  |  |  |  |  |
| < 2022 | 59,614 | 36.00 | 19.74 | 56.26 | 20 | 99.6 | < 0.001 |
| 2022 + | 5,740 | 3.40 | 0.56 | 17.96 | 8 | 98.7 | < 0.001 |
| **WHO Afro zone** |  |  |  |  |  |  |  |
| Central | 8,626 | 19.57 | 8.52 | 38.86 | 20 | 99.4 | < 0.001 |
| Western | 2,436 | 38.13 | 38.13 | 27.21 | 2 | 97.5 | < 0.001 |
| Eastern | 384 | 0.00 | 0.00 | 0.96 | 1 | - | - |
| Multi-region | 53,908 | 42.89 | 12.42 | 79.91 | 5 | 99.8 | < 0.001 |
| **Setting** |  |  |  |  |  |  |  |
| Community | 62,462 | 27.19 | 13.83 | 46.49 | 24 | 99.7 | < 0.001 |
| Hospital | 1,069 | 1.70 | 0.02 | 57.19 | 3 | 97.6 | < 0.001 |
| Online | 1,823 | 5.76 | 4.73 | 6.93 | 1 | - | - |
| **Participant** |  |  |  |  |  |  |  |
| General population | 64,417 | 23.04 | 10.64 | 42.95 | 23 | 99.7 | < 0.001 |
| General population and Healthcare worker | 162 | 21.60 | 15.94 | 28.60 | 2 | 0.0 | 0.987 |
| Healthcare worker | 775 | 5.46 | 0.04 | 89.99 | 3 | 0.0 | 0.979 |
| **Sampling** |  |  |  |  |  |  |  |
| Non-probabilistic | 54,223 | 23.26 | 11.11 | 42.35 | 23 | 99.7 | < 0.001 |
| Probabilistic | 11,131 | 11.06 | 0.74 | 67.59 | 5 | 99.8 | < 0.001 |
| **Sample size** |  |  |  |  |  |  |  |
| □ 300 | 1,251 | 18.10 | 7.49 | 37.63 | 13 | 90.6 | < 0.001 |
| 300 + | 64,103 | 23.72 | 7.60 | 54.04 | 15 | 99.8 | < 0.001 |

<sup>1</sup> Random effects model; CI: Confidence Interval; <sup>2</sup> Period before and after the mpox global outbreak <sup>3</sup> Western: Ghana and Nigeria; Eastern: Ethiopia; Central: Cameroon, Central African Republic, and Democratic Republic of Congo;

### Meta-regression analysis vaccine uptake

While univariate analysis identified study years and settings as potential factors influence mpox vaccine uptake heterogeneity, no significant study characteristics was associated with estimate heterogeneity at multivariate analysis (Table 4).

**Table 4:** Meta-regression analysis of mpox vaccination coverage in Africa, 1970-2024.

| Estimate | Moderator | Univariate |  | Multivariate |  |
| --- | --- | --- | --- | --- | --- |
| | | Unadjusted coefficient ($\beta$) | $p$-value | Adjusted coefficient ($\beta$) | $p$-value |
| Vaccination uptake |  |  |  |  |  |
| Study year | 2022+ vs. 2022 | -2.3257 | 0.005 | -2.7984 | 0.055 |
| Setting | Community | Ref | - | Ref | - |
|  | Hospital | -2.2165 | 0.010 | -0.5650 | 0.789 |
|  | Online | -1.8817 | 0.375 | -0.4735 | 0.867 |
| WHO category | Multi-region | Ref | - | Ref | - |
|  | Eastern | -6.3655 | 0.015 | -5.2509 | 0.141 |
|  | Central | -1.0410 | 0.306 | -0.8313 | 0.492 |
|  | Western | -0.1929 | 0.909 | 1.2407 | 0.606 |
| Participant | Healthcare worker | Ref | - | Ref | - |
|  | General population | 0.6822 | 0.633 | -1.2023 | 0.529 |
|  | General population and healthcare workers | 0.5218 | 0.802 | -1.3383 | 0.558 |
| Sampling | Probabilistic vs. non-probabilistic | -0.5129 | 0.639 | 0.2899 | 0.870 |
| Sample size | 300+ vs. 300 | 0.3685 | 0.657 | 0.8030 | 0.413 |

WHO: World Health Organization; Western: Ghana and Nigeria; Eastern: Ethiopia; Central: Cameroon, Central African Republic, and Democratic Republic of Congo;

### Publication bias and sensitivity test analysis

While the slight shift of the funnel plot suggested potential publication bias, Egger’s test (*p* = 0.874) and Begg’s test (*p* = 0.398) did not indicate any statistically significant bias (Additional Files 2, Supplementary Fig. 9). The sensitivity analysis showed that no single study significantly influences the pooled estimate of the vaccine acceptance rate reflecting the robustness of the finding (Additional Files 2, Supplementary Fig. 10).

The analysis reveals significant publication bias in mpox vaccine uptake studies from Africa (1970-2024), evidenced by funnel plot asymmetry (Egger’s test: *p* <0.001; Begg’s test: *p* = 0.143). The initial random-effects model showed pooled uptake of 20.94% (95%CI: 10.06-38.56) with high heterogeneity (*I*^*2*^ = 99.7%; *p* < 0.001). Trim-and-fill analysis imputed 13 missing studies (all with higher uptake estimates), adjusting the pooled rate to 71.81% (95%CI: 44.50-89.00). However, three critical limitations qualify these findings: (1) the extreme heterogeneity (*I*^*2*^ = 99.7%) suggests the adjusted estimate may overcorrect due to fundamental differences in study designs/populations rather than pure publication bias; (2) the discordant Egger’s/Begg’s tests indicate the bias may stem from small-study effects beyond just publication bias.

The robustness of vaccine uptake rate to study omission confirms the meta-analysis findings are not driven by any single study, though the consistently high heterogeneity (*I*^*2*^ = 99.7%). The Breman *et al*., 1979 study appears uniquely influential, warranting more investigation [33].

### Factors associated with mpox vaccine acceptance and uptake

The synthesis of included studies assessing vaccine acceptance and uptake identified several factors. These factors included:

**Age:** Vaccine uptake was linked to age, with individuals 15 years and older more likely to be vaccinated [34,59,60]. A higher vaccination rate was also associated with being 20 years of age or older [33,35,38], while another study found a positive association with an age exceeding 33 years [41].

**Sexe:** Vaccine uptake was higher in men [59], and gender was also associated with vaccine acceptance [25,26].

**Income:** Having a higher income has been linked to both a higher likelihood of vaccine acceptance [72] and a lower likelihood of it [26].

**Good mpox knowledge:** Higher mpox knowledge was linked to greater vaccine acceptance [16,63,64,67,70].

**Accessibility:** Free access to vaccination was associated with higher vaccine acceptance [16,24].

**Education:** Higher education levels is associated with increased vaccine acceptance [70–72]. *Risks perception:* Risk perception is also linked to greater vaccine acceptance [25,63,64,67].

**Trust in authorities and healthcare system:** Trust in governmental and healthcare institutions constituted a significant factor in vaccine adoption [25,63,72].

**Mpox vaccine trust:** Trusting the mpox vaccine to be safe is associated with higher acceptance [25,41,42,63,64].

**Mpox-related information:** Receiving mpox information from health authorities was linked to a higher likelihood of vaccine acceptance [24,41,42].

**Social norms and influences:** Positive recommendations from social circles, including peers and close contacts, were associated with a greater intention to get vaccinated [16,42].

**Previous vaccination history:** A prior vaccination history, such as having received a COVID-19 or influenza vaccine, was a significant predictor of mpox vaccine acceptance [26,63,66,67].

**Localization:** Residents in urban settings are significantly more likely to accept vaccination [25,72].

**Education and training:** Receiving specific training about mpox and vaccination increased willingness to vaccinate [16,66].

**Perceived mpox susceptibility:** The perception of being highly susceptible or at risk of mpox has been associated with a higher likelihood of intention to accept the mpox vaccine [42].

**Family history of mpox:** Families with a history of severe mpox cases demonstrated higher risk perception and greater vaccine acceptance [64].

## Discussion

This meta-analysis provides the most comprehensive synthesis of mpox vaccine acceptance and uptake in Africa, covering the period from 1970 to 2024. Framed within both continental and global perspectives, our findings reveal a moderate willingness to vaccinate (53.55%) but low recent uptake (3.4% post-2022). This underscores a persistent gap between intent and action, a gap that threatens outbreak control.

The pooled mpox vaccine acceptance rate across African studies was 53.55%, slightly lower than the global pooled estimates of 56–59.7% reported in recent systematic reviews [73,74]. Despite this relatively small difference, African acceptance exceeded averages reported for certain regions, such as Southeast Asia and the Eastern Mediterranean [13].

Marked inter-country variability was evident, with acceptance ranging from 85% in the DRC to 28.84% in Ethiopia. The higher acceptance in the DRC may be explained by its long-standing experience with mpox outbreaks, its population’s familiarity with orthopoxvirus vaccination from the smallpox eradication era, and targeted immunization programs in endemic zones [32,40,42]. In contrast, Ethiopia’s lower rates may reflect minimal direct exposure to mpox, reduced perceived personal risk, and the influence of competing public health priorities [68,75]. Similar trends have been documented globally, with elevated acceptance observed in countries recently affected by severe outbreaks, such as Nigeria and the United Kingdom during 2022–2023 [76,77].

Subregional analysis revealed the highest vaccine acceptance in Southern Africa (67.43%), with lower rates in Eastern, Central, and Western Africa. By population group, general population acceptance was moderate to high (62.46%), whereas healthcare workers (51.63%) and medical students (46.17%) demonstrated an unexpectedly lower willingness to vaccinate. This finding echoes results from Ebola and COVID-19 vaccination campaigns, where occupational exposure alone did not consistently lead to higher uptake [16,25,63]. Contributing factors may include vaccine safety concerns, limited mpox-specific training, and exclusion from priority vaccination lists [25,78].

Beyond acceptance rates, our analysis reveals significant disparities in actual mpox vaccine coverage across Africa. The most notable temporal pattern was the sharp drop in uptake from 36.0% before 2022 to just 3.4% post-2022. This decline is likely due to multiple factors, including limited vaccine availability, dependence on donor-driven allocations, and the absence of sustained, large-scale vaccination campaigns after the initial outbreak response. The WHO’s 2023 decision to declare mpox no longer a Public Health Emergency of International Concern may have further diminished perceived urgency, contributing to reduced demand and political prioritization [79].

The mpox vaccination coverage in Africa was 20.94% (95% CI: 10.06-38.56). Our findings were non-significantly lower than observed in global pooled estimate of 30.9% (95% CI, 21.0– 41.7%) [13]. This highlight the geographiscal disparity in vaccine delivery across various regions globally. Moreover, The gap between vaccine acceptance and actual uptake was particularly pronounced among healthcare workers (5.5%) and hospital-based populations (1.7%). In Eastern Africa, the only available study reported 0% coverage. This significant mismatch echoes patterns seen with Ebola and COVID-19 vaccination efforts in Africa, where acceptance often exceeded 60%, but coverage lagged due to logistical challenges, inequitable distribution, and inconsistent risk communication [78,80,81]. These persistent barriers highlight the urgent need to strengthen vaccine supply chains, enhance targeted outreach, and embed mpox vaccination into routine immunization programs.

Historically, smallpox vaccination campaigns in Africa achieved coverage exceeding 90%[40]. This underscores the feasibility of high uptake and highlights the extent of the current gaps. Smallpox eradication succeeded due to ample vaccine supply, efficient delivery systems, and a high public threat perception. In contrast, the current mpox landscape is marked by a breakdown of the orthopoxvirus vaccination infrastructure, diminished public familiarity with the disease, and competition from other immunization priorities [40].

This study adds new knowledge by quantifying the magnitude of the acceptance-uptake gap for mpox vaccines in Africa. It identifies greater inter-country variability compared to global trends, documents a sharp decline in uptake post-2022, and reveals unexpectedly low acceptance among healthcare workers. These findings suggest that strategies focused solely on demand generation are insufficient. Instead, a holistic approach addressing both vaccine supply and systemic delivery constraints is required.

The gap between vaccine acceptance and actual uptake was particularly pronounced among healthcare workers (5.5%) and hospital-based populations (1.7%). In Eastern Africa, the only available study reported 0% coverage. This significant mismatch echoes patterns seen with Ebola and COVID-19 vaccination efforts in Africa, where acceptance often exceeded 60%, but coverage lagged due to logistical challenges, inequitable distribution, and inconsistent risk communication [82–84]. These persistent barriers highlight the urgent need to strengthen vaccine supply chains, enhance targeted outreach, and embed mpox vaccination into routine immunization programs in countries facing recurrent outbreaks.

Historically, smallpox vaccination campaigns in Africa achieved coverage exceeding 90% [40]. This underscores the feasibility of high uptake and highlights the extent of the current gaps. Smallpox eradication succeeded due to ample vaccine supply, efficient delivery systems, and a high public threat perception. In contrast, the current mpox landscape is marked by a breakdown of the orthopoxvirus vaccination infrastructure, diminished public familiarity with the disease, and competition from other immunization priorities [40].

This study adds new knowledge by quantifying the magnitude of the acceptance-uptake gap for mpox vaccines in Africa. It identifies greater inter-country variability compared to global trends, documents a sharp decline in uptake post-2022, and reveals unexpectedly low acceptance among healthcare workers. These findings suggest that strategies focused solely on demand generation are insufficient. Instead, a holistic approach addressing both vaccine supply and systemic delivery constraints is required.

Furthermore, this study showed that mpox vaccine acceptance and uptake in Africa are influenced by a complex interplay of sociodemographic, psychosocial, and structural factors. Age and sex emerged as significant determinants. Older individuals were more likely to accept vaccination than younger people [33–35,60,82–84], a finding that corroborates previous COVID-19 vaccine studies where older adults reported higher willingness due to a greater perceived vulnerability [85].

Evidence on gender-based acceptance was mixed. Some studies found higher uptake among men [16], while others identified greater acceptance among women [40,43]. This variability mirrors COVID-19 research in Africa, where male acceptance was sometimes higher due to occupational exposure, while female hesitancy was linked to concerns about fertility and vaccine safety [86,87].

In addition to sociodemographic factors, socioeconomic and educational factors also shaped vaccine acceptance in our review. Higher education consistently predicted willingness to vaccinate [70–72], which corroborates previous studies showing that health literacy enhances vaccine confidence [88]. However, the effects of income were contradictory: higher income was associated with both greater [72] and lower acceptance [26]. These inconsistencies likely reflect contextual differences, as wealth may improve access in some settings but coincide with skepticism of government or donor-driven campaigns in others [89]. Beyond formal education, knowledge about mpox, risk perception, and perceived susceptibility were strongly associated with acceptance [16,63,63,67,70]. This aligns with the Health Belief Model, where perceived risk drives preventive health behavior [90]. Additionally, training interventions [16,66] boosted willingness, echoing evidence from Ebola and COVID-19 vaccination programs in Africa where targeted education improved uptake [91].

Trust and information emerged as central themes. Confidence in health authorities and the healthcare system [25,72,92], and trust in the vaccine’s safety [25,41,92,92,93], were critical drivers, consistent with global evidence that institutional trust underpins vaccine confidence [94]. Conversely, mistrust has repeatedly undermined campaigns during Ebola and and COVID-19 outbreaks [95]. Receiving information from health authorities increased acceptance [24,41,93], while social influences such as peer and family recommendations also shaped decisions [16,93]. A prior vaccination history, including for COVID-19 or influenza, predicted acceptance [26,66,67,92], suggesting that established immunization habits carry over to new vaccines [96]. Structural barriers were equally important: free provision [16,24] and urban residence [25,72] increased uptake, while rural populations faced greater challenges, consistent with long-standing inequities in African immunization programs [97]. Finally, family history of mpox increased both risk perception and acceptance [92], mirroring Ebola outbreaks where personal experience heightened demand [98].

Taken together, these findings show that improving mpox vaccine uptake in Africa requires multi-level interventions that address structural inequities in access, tailor education and communication for different sociodemographic groups, build trust through transparent messaging, and leverage prior vaccination experiences. The complex interplay of individual, social, and systemic factors underscores the need for context-specific strategies to ensure that acceptance translates into equitable uptake across the continent.

Importantly, this study adds new knowledge by quantifying the magnitude of the acceptance–uptake gap for mpox vaccines in Africa, identifying greater inter-country variability compared with global trends, documenting a post-2022 collapse in uptake, and revealing unexpectedly low acceptance among healthcare workers. These findings suggest that strategies focusing solely on demand generation may be insufficient. Instead, holistic approaches that simultaneously address vaccine supply, equitable distribution, and systemic delivery challenges are needed to effectively convert stated willingness into completed vaccinations.

### Strength and limitations

This systematic review and meta-analysis provide a comprehensive synthesis of Mpox vaccination trends in Africa over five decades, offering valuable insights into temporal patterns, geographic disparities, and implementation gaps through robust analysis. However, this review has several limitations. The small sample sizes in some subgroup analyses resulted in wide confidence intervals, limiting the precision of the findings. The predominance of non-probabilistic surveillance data might limit the inference. In addition, the vaccination status relied primarily on individual reports or the presence of a scar of vaccination instead of the presentation of a vaccination card testifying to the reception of the vaccine.

## Conclusions

Mpox vaccine acceptance in Africa remains moderate overall but highly heterogeneous across countries, regions, and population groups. While willingness to vaccinate often approaches or exceeds global averages, actual coverage is markedly lower, with a sharp post-2022 decline highlighting fragility in vaccine supply and delivery systems. The pronounced acceptance–uptake gap, especially among healthcare workers and in high-risk settings, underscores that generating demand is only part of the solution. Achieving effective mpox vaccination coverage will require sustained political commitment, equitable and reliable vaccine supply chains, targeted communication strategies, and integration into routine immunization platforms.

## Supporting information

Addition Files 2

Addition Files 3

## Data Availability

All data generated or analyzed during this study are included in this published article and supplemental material.

## Abbreviations

CI: Confidence interval
DRC: Democratic Republic of Congo
MeSH: Medical subject headings
mpox: Monkeypox
PRISMA: Preferred reporting items for systematic reviews and meta-analysis

## Declarations

### Ethical approval and consent to participate

Not applicable. *Consent for publication:* Not applicable.

### Availability of data and materials

The sources of data supporting this systematic review are available in the reference. All data generated or analyzed during this study are included in this published article and supplemental material.

### Author contributions

F.Z.L.C. conceived the original idea of the study. F.Z.L.C and M.N.T. conducted the literature search. F.Z.L.C., R.T., A.C. and A.A.N. selected the studies, extracted the relevant information, critically assessed included reports and synthesized the data. F.Z.L.C. performed the analyses. F.Z.L.C., R.T. A.C. and A.A.N. wrote the first draft of the manuscript. All authors critically reviewed and revised successive drafts of the manuscript. All authors read and approved the final manuscript.

## Acknowledgements

None.

## References

1. Islam MA, Mumin J, Haque MM, Haque MdA, Khan A, Bhattacharya P, et al. Monkeypox virus (MPXV): A Brief account of global spread, epidemiology, virology, clinical features, pathogenesis, and therapeutic interventions. Infect Med. 2023;2(4):262–262.

2. Mpox (monkeypox). WHO | Regional Office for Africa, Brazzaville. 2025. https://www.afro.who.int/health-topics/mpox-monkeypox. Accessed: 2025 Sep 2.

3. Subissi L, Mitjà O. Rising mpox trends in DR Congo: the neglected spread of an epidemic. The Lancet. 2025;405(10476):358–60.

4. Cheuyem FZL, Zemsi A, Ndungo JH, Achangwa C, Takpando-le-grand DR, Goupeyou-Youmsi J, et al. Mpox severity and mortality in the most endemic focus in africa: a systematic review and meta-analysis (1970-2024). medRxiv; 2025. doi: 10.1101/2025.04.07.25325410.

5. Akingbola A, Adegbesan CA, Adewole O, Idahor C, Odukoya T, Nwaeze E, et al. Understanding the resurgence of mpox: key drivers and lessons from recent outbreaks in Africa. Trop Med Health. 2025;53(1):47.

6. Djuicy DD, Sadeuh-Mba SA, Bilounga CN, Yonga MG, Tchatchueng-Mbougua JB, Essima GD, et al. Concurrent Clade I and Clade II Monkeypox Virus Circulation, Cameroon, 1979–2022. Emerg Infect Dis. 2024;30(3):433–433.

7. Ebede SO, Orabueze IN, Maduakor UC, Nwafia IN, Ohanu ME. Recurrent Mpox: divergent virulent clades and the urgent need for strategic measures including novel vaccine development to sustain global health security. BMC Infect Dis. 2025;25(1):536.

8. Clade I Mpox Outbreak Originating in Central Africa. CDC, Atlanta. 2025. https://www.cdc.gov/mpox/outbreaks/2023/index.html. Accessed: 2025 Jul 1.

9. Hajjo R, Abusara OH, Sabbah DA, Bardaweel SK. Advancing the understanding and management of Mpox: insights into epidemiology, disease pathways, prevention, and therapeutic strategies. BMC Infect Dis. 2025;25(1):529.

10. Lâm S, Masudi SP, Nguyen HTT, Grace D. Preventing mpox at its source: Using food safety and One Health strategies to address bushmeat practices. BMC Glob Public Health. 2024;2(1):69.

11. Preventing Mpox. CDC, Atlanta. 2024. https://www.cdc.gov/mpox/prevention/index.html. Accessed: 2025 Sep 2.

12. Multi-country outbreak of mpox, External situation report #53 - 29 May 2025. WHO, Geneva. 2025. https://www.who.int/publications/m/item/multi-country-outbreak-of-mpox--external-situation-report--5329-may-2025. Accessed: 2025 Jul 1.

13. Sulaiman SK, Isma’il Tsiga-Ahmed F, Musa MS, Makama BT, Sulaiman AK, Abdulaziz TB. Global prevalence and correlates of mpox vaccine acceptance and uptake: a systematic review and meta-analysis. Commun Med. 2024;4(1):136.

14. Sah R, Humayun M, Baig E, Farooq M, Hussain HG, Shahid MU, et al. FDA’s authorized “JYNNEOS” vaccine for counteracting monkeypox global public health emergency; an update - Correspondence. Int J Surg Lond Engl. 2022; 107:106971.

15. Chakraborty C, Bhattacharya M, Ranjan Sharma A, Dhama K. Monkeypox virus vaccine evolution and global preparedness for vaccination. Int Immunopharmacol. 2022; 113:109346.

16. Ghazy RM, Hussein M, Abdu SMM, El-sayed Ellakwa D, Tolba MM, Youssef N, et al. The intention of Egyptian healthcare workers to take the monkeypox vaccine: is urgent action required? BMC Health Serv Res. 2024;24(1):1204.

17. Alie MS, Abebe GF, Negesse Y, Adugna A, Girma D. Vaccine hesitancy in context of COVID-19 in East Africa: systematic review and meta-analysis. BMC Public Health. 2024;24(1):2796.

18. Uğrak U, Aksungur A, Akyüz S, Şen H, Seyhan F. Understanding the rise of vaccine refusal: perceptions, fears, and influences. BMC Public Health. 2025;25(1):2574.

19. Cheuyem FZL, Amani A, Nkodo ICA, Boukeng LBK, Edzamba MF, Nouko A, et al. COVID-19 vaccine acceptance and hesitancy in Cameroon: a systematic review and meta-analysis. BMC Public Health. 2025;25(1):1035.

20. Cheuyem FZL, Amani A, Achangwa C, Ajong BN, Minkandi CA, Zeh MMMK, et al. COVID-19 vaccine uptake and its determinants in Cameroon: a systematic review and meta-analysis (2021–2024). BMC Infect Dis. 2025;25(1):525.

21. Cheuyem FZL, Lyonga EE, Kamga HG, Mbopi-Keou FX, Takougang I. Needlestick and Sharp Injuries and Hepatitis B Vaccination among Healthcare Workers: A Cross-Sectional Study in Six District Hospitals in Yaounde (Cameroon). J Community Med Public Health. 2023;7(3):1–9.

22. Takougang I, Cheuyem FZL, Lyonga EE, Ndungo JH, Mbopi-Keou FX. Observance of Standard Precautions for Infection Prevention in The Covid-19 Era: A Cross-Sectional Study in Six District Hospitals in Yaounde, Cameroon. Am J Biomed Sci Res. 2023;19(5):590–590.

23. Amani A, Mossus T, Cheuyem FZL, Bilounga C, Mikamb P, Basseguin Atchou J, et al. Gender and COVID-19 Vaccine Disparities in Cameroon. COVID. 2022;2(12):1715–30.

24. Petrichko S, Kindrachuk J, Nkamba D, Halbrook M, Merritt S, Kalengi H, et al. Mpox Vaccine Acceptance, Democratic Republic of the Congo. Emerg Infect Dis. 2024;30(12):2614–2614.

25. Fetensa G, Tolossa T, Besho M, Yadesa G, Gugsa J, Tufa DG, et al. Willingness to take Mpox vaccine and associated factors among health professionals in Ethiopia: A cross-sectional study. Vaccine. 2025; 49:126822.

26. Braimah JA, Achore M, Dery F, Ayanore MA, Bisung E, Kuuire V. Do self-rated health and previous vaccine uptake influence the willingness to accept MPOX vaccine during a public health emergency of concern? A cross-sectional study. Robinson J, editor. PLOS Glob Public Health. 2024;4(8): e0003564.

27. Mpox Vaccination. Africa CDC. https://africacdc.org/mpox/mpox-vaccination/. Accessed: 2025 Sep 2.

28. Elechi KW, Toluwalashe S, Oyelude A, Okechukwu CI, Obed M, Okoli GC, et al. Building Bridges: The Role of Effective Community Engagement Strategies for Mpox Prevention and Response. Afro-Egypt J Infect Endem Dis. 2025;15(3):226–226.

29. Mambo SB, Nja GME, Bunu UO, Bizimana GN, M’yisa Makelele A, Sonia FD, et al. Promoting risk communication and community engagement during Mpox outbreak in fragile conflict zones of Eastern DRC. One Health. 2025; 20:101012.

30. Mithi VS. Mpox in Africa: funding cuts and delayed global actions fuelling new epicentres. Lancet Glob Health. 2025;13(9): e1512–3.

31. Kangbai JB, Sesay U, Vickos U, Kagbanda F, Fallah MP, Osborne A. Mpox in Africa: What we know and what is still lacking. PLoS Negl Trop Dis. 2025;19(6): e0013148.

32. Ladnyj ID, Ziegler P, Kima E. A human infection caused by monkeypox virus in Basankusu Territory, Democratic Republic of the Congo. Bull World Health Organ. 1972;46(5):593–7.

33. Breman JG, Kalisa-Ruti Steniowski MV, Zanotto E, Gromyko AI, Arita I. Human monkeypox, 1970-79. Bull World Health Organ. 1980;58(2):165–165.

34. Mutombo M, Jezek Z, Arita I, Jezek Z. Human monkeypox transmitted by a chimpanzee in a tropical rain-forest area of Zaire. Lancet Lond Engl. 1983;321(8327):735–7.

35. Jezek Z, Marennikova SS, Mutumbo M, Nakano JH, Paluku KM, Szczeniowski M. Human Monkeypox: A Study of 2,510 Contacts of 214 Patients. J Infect Dis. 1986;154(4):551–551.

36. Jezek Z, Szczeniowski M, Paluku KM, Mutombo M. Human Monkeypox: Clinical Features of 282 Patients. J Infect Dis. 1987;156(2):293–8.

37. Mwanba PT, Tshioko KF, Moudi A, Mukinda V, Mwema GN, Messinger D, et al. Human monkeypox in Kasaï Oriental, Zaire (1996-1997). Eurosurveillance. 1997;2(5):33–5.

38. Hutin YJF, Williams RJ, Malfait P, Pebody R, Loparev VN, Ropp SL, et al. Outbreak of Human Monkeypox, Democratic Republic of Congo, 1996 to 1997. Emerg Infect Dis. 2001;7(3):434–434.

39. Aplogan A, Mangindula V, Muamba P, Mwema G, Okito L, Pebody R, et al. Human monkeypox -- Kasai Oriental, Democratic Republic of Congo, February 1996-October 1997. MMWR Morb Mortal Wkly Rep. 1997;46(49):1168–1168.

40. Rimoin AW, Mulembakani PM, Johnston SC, Lloyd Smith JO, Kisalu NK, Kinkela TL, et al. Major increase in human monkeypox incidence 30 years after smallpox vaccination campaigns cease in the Democratic Republic of Congo. Proc Natl Acad Sci. 2010;107(37):16262–16262.

41. Nolen LD, Osadebe L, Katomba J, Likofata J, Mukadi D, Monroe B, et al. Extended Human-to-Human Transmission during a Monkeypox Outbreak in the Democratic Republic of the Congo. Emerg Infect Dis. 2016;22(6):1014–21.

42. Petersen BW, Kabamba J, McCollum AM, Lushima RS, Wemakoy EO, Muyembe Tamfum JJ, et al. Vaccinating against monkeypox in the Democratic Republic of the Congo. Antiviral Res. 2019; 162:171–7.

43. McCollum AM, Nakazawa Y, Ndongala GM, Pukuta E, Karhemere S, Lushima RS, et al. Human Monkeypox in the Kivus, a Conflict Region of the Democratic Republic of the Congo. Am Soc Trop Med Hyg. 2015;93(4):718–1.

44. Whitehouse ER, Bonwitt J, Hughes CM, Lushima RS, Likafi T, Nguete B, et al. Clinical and Epidemiological Findings from Enhanced Monkeypox Surveillance in Tshuapa Province, Democratic Republic of the Congo During 2011–2015. J Infect Dis. 2021;223(11):1870–1870.

45. Mande G, Akonda I, Weggheleire AD, Brosius I, Liesenborghs L, Bottieau E, et al. Enhanced surveillance of monkeypox in Bas-Uélé, Democratic Republic of Congo: the limitations of symptom-based case definitions. Int J Infect Dis. 2022; 122:647–55.

46. Brosius I, Vakaniaki EH, Mukari G, Munganga P, Tshomba JC, Vos ED, et al. Epidemiological and clinical features of mpox during the clade Ib outbreak in South Kivu, Democratic Republic of the Congo: a prospective cohort study. The Lancet. 2025;405(10478):547–59.

47. Kombozi YDMerci, Singa Valentin. B, Ley. BL, Antoine. ET, Guild. AK, Lwanga. K, et al. Mpox: epidemiological profile and factors associated with its emergence in the isangi territory, Democratic Republic of Congo. Int J Appl Sci Eng Rev. 2024;5(5):27–27.

48. Vakaniaki EH, Kacita C, Kinganda-Lusamaki E, O’Toole Á, Wawina-Bokalanga T, Mukadi-Bamuleka D, et al. Sustained human outbreak of a new MPXV clade I lineage in eastern Democratic Republic of the Congo. Nat Med. 2024;30(10):2791–5.

49. Moher D, Liberati A, Tetzlaff J, Altman DG, Group TP. Preferred Reporting Items for Systematic Reviews and Meta-Analyses: The PRISMA Statement. PLOS Med. 2009;6(7):e1000097.

50. Cheuyem FZL, Njoh AA, Achangwa C, Kistner O, Tchamani R, Goupeyou-Youmsi J, et al. Mpox Vaccination Acceptance and Uptake in Africa (1970–2024): A systematic Review and Metanalysis. PROSPERO 2025 CRD420251126033. https://www.crd.york.ac.uk/PROSPERO/view/CRD420251126033. Accessed: 2025 Sep 17.

51. Munn Z, Stern C, Aromataris E, Lockwood C, Jordan Z. What kind of systematic review should I conduct? A proposed typology and guidance for systematic reviewers in the medical and health sciences. BMC Med Res Methodol. 2018;18(1):5.

52. JBI Critical Appraisal Tools. The Joanna Briggs Institute, Adelaide. 2017. https://jbi.global/critical-appraisal-tools. Accessed: 2024 Dec 3.

53. ESPEN. WHO | Regional Office for Africa. 2023. https://espen.afro.who.int/maps-data/regional-maps-data/afro. Accessed: 2025 Jul 14.

54. Stijnen T, Hamza TH, Ozdemir P. Random effects meta-analysis of event outcome in the framework of the generalized linear mixed model with applications in sparse data. Stat Med. 2010;29(29):3046–67.

55. R Core Team. R: A Language and Environment for Statistical Computing. R Foundation for Statistical Computing, Vienna, Austria. 2024. https://www.R-project.org/. Accessed: 2024 May 30.

56. Create your own Custom Map. MapChart. 2025 [Accessed: 2025 Aug 27]. https://mapchart.net/index.html

57. Egger M, Davey Smith G, Schneider M, Minder C. Bias in meta-analysis detected by a simple, graphical test. BMJ. 1997;315(7109):629–629.

58. Begg CB, Mazumdar M. Operating characteristics of a rank correlation test for publication bias. Biometrics. 1994 Dec;50(4):1088.

59. Henderson RH, Davis H, Eddins DL, Foege WH. Assessment of vaccination coverage, vaccination scar rates, and smallpox scarring in five areas of West Africa. Bull World Health Organ. 1973;48(2):183.

60. Arita I, Jezek Z, Khodakevich L, Ruti K. Human monkeypox: a newly emerged orthopoxvirus zoonosis in the tropical rain forests of Africa. Am J Trop Med Hyg. 1985;34(4):781–781.

61. Kalthan E, Dondo-Fongbia JP, Yambele S, Dieu-Creer LR, Zepio R, Pamatika CM. Epidémie de 12 cas de maladie à virus monkeypox dans le district de Bangassou en République Centrafricaine en décembre 2015. Bull Société Pathol Exot. 2016;109(5):358–63.

62. Besombes C, Gonofio E, Konamna X, Selekon B, Gessain A, Berthet N, et al. Intrafamily Transmission of Monkeypox Virus, Central African Republic, 2018. Emerg Infect Dis. 2019;25(8):1602–1602.

63. Lounis M, Bencherit D, Abdelhadi S. Knowledge and awareness of Algerian healthcare workers about human monkeypox and their attitude toward its vaccination: An online cross-sectional survey. Vacunas. 2023;24(2):122–7.

64. Lounis M, Hamimes A, Dahmani A. Assessment of Monkeypox (MPOX) Knowledge and Vaccination Intention among Health and Life Sciences Students in Algeria: A Cross-Sectional Study. Infect Dis Rep. 2024;16(2):170–170.

65. Nganmegni FRP, Ngo JLL, Essomba RG, Nguwoh PS, Metomb FS, Epee E, et al. Burden and predictors of M-pox suspected cases in a rural setting of Cameroon: implications for developing countries. J Infect Dev Ctries. 2024;18(11):1756–1756.

66. Amer FA, Nofal HA, Gebriel MG, Bedawy AM, Allam AA, Khalil HES, et al. Grasping knowledge, attitude, and perception towards monkeypox among healthcare workers and medical students: an Egyptian cross-sectional study. Front Cell Infect Microbiol. 2024; 14:1339352.

67. Hussein M, Siddiq A, Ismail HM, Mansy N, Ellakwa DES, Nassif M, et al. Cross-Country Discrepancies in Monkeypox Vaccine Hesitancy Among Postgraduate and Undergraduate Medical Students. Disaster Med Public Health Prep. 2024;18: e82.

68. Aynalem Z, Abate M, Meseret F, Muhamed A, Abebe G, Adal A, et al. Knowledge, Attitude and Associated Factors of Monkeypox Infection Among Healthcare Workers in Injibara General Hospital, Northwest Ethiopia. J Multidiscip Healthc. 2024; 17:1159–73.

69. Du M, Deng J, Yan W, Liu M, Liang W, Niu B, et al. Mpox vaccination hesitancy, previous immunisation coverage, and vaccination readiness in the African region: a multinational survey. eClinicalMedicine. 2025; 80:103047.

70. Kiros T, Erkihun M, Wondmagegn M, Almaw A, Assefa A, Berhan A, et al. Assessment of Knowledge, Attitude, and Associated Factors of Mpox Among Healthcare Professionals at Debre Tabor Comprehensive Specialized Hospital, Northwest Ethiopia, 2024: A Cross-Sectional Study. Health Sci Rep. 2025;8(1): e70371.

71. Mutua PM, Gicheru MM, Mutiso J, Serem E. A cross-sectional study of Mpox Knowledge, Attitudes toward Mpox Vaccination and Mpox Vaccine Hesitancy Among Teachers in Kenya. In Review; 2024. https://www.researchsquare.com/article/rs-5458486/v1. Accessed: 2025 Jul 6.

72. Lawrence A. Socio-Demographic Factors Influencing Monkeypox Vaccination Intentions Among Healthcare Workers and the General Population of Makurdi, Nigeria: A Cross-Sectional Study. Cureus. 2024;16(10): e71828.

73. Mektebi A, Elsaid M, Yadav T, Abdallh F, Assker M, Siddiq A, et al. Mpox vaccine acceptance among healthcare workers: a systematic review and meta-analysis. BMC Public Health. 2024;24(1):4.

74. Yappalparvi A, Gaidhane S, Padmapriya G, Kaur I, Lal M, Iqbal S, et al. Prevalence of Mpox Vaccine Acceptance Among Students: A Systematic Review and Meta-Analysis. Vaccines. 2025;13(2):183.

75. Ayele HS, Mengesha AK, Geremew GW, Lakew AA, Alemayehu TT, Getachew D, et al. Assessment of Knowledge, Attitude, and Associated Factors towards Monkeypox Infection among residents at Bahir Dar city, Northwest Ethiopia, 2024. Community based cross-sectional study. SAGE Open Nurs. 2025; 11:23779608251352392.

76. Sulaiman SK, Isma’il Tsiga-Ahmed F, Musa MS, Makama BT, Sulaiman AK, Abdulaziz TB. Global prevalence and correlates of mpox vaccine acceptance and uptake: a systematic review and meta-analysis. Commun Med. 2024;4(1):136.

77. Karapinar A, AkdaĞ D, GÖkengİn D. Awareness and acceptability of monkeypox vaccine in men who have sex with men. Turk J Med Sci. 53(5):1136–43.

78. Solís Arce JS, Warren SS, Meriggi NF, Scacco A, McMurry N, Voors M, et al. COVID-19 vaccine acceptance and hesitancy in low- and middle-income countries. Nat Med. 2021;27(8):1385–1385.

79. Sarker R, Roknuzzaman ASM, Shahriar M, Bhuiyan MA, Islam MdR. The WHO has ended public health emergency of international concern for mpox: assessment of upside and downside of this decision. Int J Surg Lond Engl. 2023;109(10):3238–3238.

80. Amani A, Bene ACM, Lungoyo CL, Mukoka AK, Cheuyem FZL, Mpinganjira S, et al. The Struggle to Vaccinate: Unveiling the Reality of the first year of Covid-19 Vaccination in the Democratic Republic of Congo. medRxiv; 2024. doi: 10.1101/2024.01.03.24300795.

81. Kpanake L, Sorum PC, Mullet É. Willingness to get vaccinated against Ebola: A mapping of Guinean people positions. Hum Vaccines Immunother. 2018;14(10):2391–2391.

82. Henderson RH, Davis H, Eddins DL, Foege WH. Assessment of vaccination coverage, vaccination scar rates, and smallpox scarring in five areas of West Africa. Bull World Health Organ. 1973;48(2):183–183.

83. Hutin YJ, Williams RJ, Malfait P, Pebody R, Loparev VN, Ropp SL, et al. Outbreak of human monkeypox, Democratic Republic of Congo, 1996 to 1997. Emerg Infect Dis. 2001;7(3):434–434.

84. Nolen LD, Osadebe L, Katomba J, Likofata J, Mukadi D, Monroe B, et al. Extended Human-to-Human Transmission during a Monkeypox Outbreak in the Democratic Republic of the Congo. Emerg Infect Dis. 2016;22(6):1014–1014.

85. Troiano G, Nardi A. Vaccine hesitancy in the era of COVID-19. Public Health. 2021; 194:245–51.

86. Solís Arce JS, Warren SS, Meriggi NF, Scacco A, McMurry N, Voors M, et al. COVID-19 vaccine acceptance and hesitancy in low- and middle-income countries. Nat Med. 2021;27(8):1385–94.

87. AlShurman BA, Khan AF, Mac C, Majeed M, Butt ZA. What Demographic, Social, and Contextual Factors Influence the Intention to Use COVID-19 Vaccines: A Scoping Review. Int J Environ Res Public Health. 2021;18(17):9342.

88. Bish A, Yardley L, Nicoll A, Michie S. Factors associated with uptake of vaccination against pandemic influenza: A systematic review. Vaccine. 2011;29(38):6472–84.

89. Dubé E, Laberge C, Guay M, Bramadat P, Roy R, Bettinger JA. Vaccine hesitancy. Hum Vaccines Immunother. 2013;9(8):1763–1763.

90. Rosenstock IM. Historical Origins of the Health Belief Model. Health Educ Monogr. 1974;2(4):328–328.

91. Sallam M. COVID-19 Vaccine Hesitancy Worldwide: A Concise Systematic Review of Vaccine Acceptance Rates. Vaccines. 2021;9(2):160.

92. Lounis M, Bencherit D, Abdelhadi S. Knowledge and awareness of Algerian healthcare workers about human monkeypox and their attitude toward its vaccination: An online cross-sectional survey. Vacunas. 2023;24(2):122–7.

93. Petersen BW, Kabamba J, McCollum AM, Lushima RS, Wemakoy EO, Muyembe Tamfum JJ, et al. Vaccinating against monkeypox in the Democratic Republic of the Congo. Antiviral Res. 2019; 162:171–7.

94. (PDF) State of vaccine confidence in the European Union in 2018. ResearchGate. https://www.researchgate.net/publication. Accessed: 2025 Aug 22.

95. Cooper S, Rooyen H van, Wiysonge CS. COVID-19 vaccine hesitancy in South Africa: how can we maximize uptake of COVID-19 vaccines? Expert Rev Vaccines. 2021. doi: 10.1080/14760584.2021.1949291.

96. Schmid P, Rauber D, Betsch C, Lidolt G, Denker ML. Barriers of Influenza Vaccination Intention and Behavior – A Systematic Review of Influenza Vaccine Hesitancy, 2005 – 2016. PLoS ONE. 2017;12(1): e0170550.

97. Mihigo R, Okeibunor J, Anya B, Mkanda P, Zawaira F. Challenges of immunization in the African Region. Pan Afr Med J. 2017;27(Suppl 3):12.

98. Ditekemena JD, Nkamba DM, Mutwadi A, Mavoko HM, Siewe Fodjo JN, Luhata C, et al. COVID-19 Vaccine Acceptance in the Democratic Republic of Congo: A Cross-Sectional Survey. Vaccines. 2021;9(2):153.

