## Additional Files 1 for "Mpox vaccine acceptance and uptake in Africa: a systematic review and meta-analysis (1970-2024)"

**Supplementary Table 1** Characteristics of studies assessing vaccination acceptance and uptake in Africa, 1970-2024

| **Author** | **Reference** | **Study year** | **Country** | **Setting** | **Participant** | **Sampling** | **Outcome assessed** | **Risk bias** |
| --- | --- | --- | --- | --- | --- | --- | --- | --- |
| Ladnyj,1970 | [1] | 1970 | DRC | Community | General population | Non-probabilistic | VU | Low |
| Henderson,1973 | [2] | 1973 | Multi-country | Community | General population | Probabilistic | VU | Low |
| Breman,1979 | [3] | 1979 | Multi-country | Community | General population | Non-probabilistic | VU | Low |
| Mutombo,1982 | [4] | 1982 | DRC | Community | General population | Non-probabilistic | VU | Low |
| Arita,1983 | [5] | 1983 | Multi-country | Community | General population | Non-probabilistic | VU | Low |
| Jezek,1985 | [6] | 1985 | DRC | Community | General population | Non-probabilistic | VU | Low |
| Jezek,1984 | [7] | 1986 | DRC | Community | General population | Non-probabilistic | VU | Low |
| Mwamba,1997 | [8] | 1997 | DRC | Community | General population | Non-probabilistic | VU | Low |
| Hutin,1997 | [9] | 1997 | DRC | Community | General population | Non-probabilistic | VU | Low |
| Aplogan,1997 | [10] | 1997 | DRC | Community | General population | Non-probabilistic | VU | Low |
| Rimoin,2007 | [11] | 2007 | DRC | Community | General population | Non-probabilistic | VU | Low |
| Nolen,2013 | [12] | 2013 | DRC | Community | General population and HCWs | Non-probabilistic | VA and VU | Low |
| Petersen,2014 | [13] | 2014 | DRC | Community | HCWs | Non-probabilistic | VA and VU | Low |
| McCollum,2014 | [14] | 2014 | DRC | Community | General population | Non-probabilistic | VU | Low |
| Whitehouse,2015 | [15] | 2015 | DRC | Community | General population | Non-probabilistic | VU | Low |
| Kalthan,2016 | [16] | 2016 | Central African Republic | Community | General population | Non-probabilistic | VU | Low |
| Besombes,2018 | [17] | 2018 | Central African Republic | Community | General population and HCWs | Non-probabilistic | VU | Moderate |
| Mande,2019 | [18] | 2019 | DRC | Community | General population | Non-probabilistic | VU | Low |
| Lounis,2022 | [19] | 2022 | Algeria | Online | HCWs | Non-probabilistic | VA | Moderate |
| Nganmegni,2022 | [20] | 2022 | Cameroon | Community | General population | Non-probabilistic | VU | Low |
| Amer,2022 | [21] | 2022 | Egypt | Hospital | HCWs | Non-probabilistic | VA | Low |
| Hussein,2022 | [22] | 2022 | Multi-country | Online | Medical students | Non-probabilistic | VA | Low |
| Aynalem,2022 | [23] | 2022 | Ethiopia | Hospital | HCWs | Probabilistic | VA | Low |
| Ghazy,2022 | [24] | 2022 | Ghana | Online | HCWs | Non-probabilistic | VA | Low |
| Braimah,2023 | [25] | 2023 | Ghana | Community | General population | Probabilistic | VA and VU | Low |
| Lounis,2023 | [26] | 2023 | Algeria | Online | Medical students | Non-probabilistic | VA | Moderate |
| Brosius,2024 | [27] | 2024 | DRC | Hospital | General population | Non-probabilistic | VU | Moderate |
| Petrichko,2024 | [28] | 2024 | DRC | Community | General population | Non-probabilistic | VA | Moderate |
| Du,2024 | [29] | 2024 | Multi-country | Online | General population | Non-probabilistic | VA and VU | Low |
| Fetensa,2024 | [30] | 2024 | Ethiopia | Online | HCWs | Probabilistic | VA | Low |
| Kiros,2024 | [31] | 2024 | Ethiopia | Hospital | HCWs | Probabilistic | VA and VU | Low |
| Mutua,2024 | [32] | 2024 | Kenya | Online | Teachers | Non-probabilistic | VA | Low |
| Lawrence,2024 | [33] | 2024 | Nigeria | Hospital | HCWs | Probabilistic | VA and VU | Low |
| Kombozi,2024 | [34] | 2024 | DRC | Community | General population | Probabilistic | VU | Low |
| Vakaniaki,2024 | [35] | 2024 | DRC | Community | General population | Non-probabilistic | VU | Low |
| DRC: Democratic Republic of Congo; CS: Cross-sectional study; HCW: Healthcare worker; VU: Vaccine uptake; VA: Vaccine acceptance | | | | | | | | |

**Supplementary Table 2** Searching strategy and results by database

| Search Term | Database | Results |
| --- | --- | --- |
| (Mpox) OR (mpox) OR (Monkeypox) AND (vaccine) OR (vaccination) OR (acceptance) AND (African countries) | PubMed | 2,245 |
|  | Web of Science | 1,128 |
|  | Scopus | 3,174 |
|  | CINAHL | 5,17 |
|  | Embase | 1,224 |
|  | ScienceDirect | 1,207 |
|  | African Journals Online (AJOL) | 108 |
