## Supplementary material for "Mpox vaccine acceptance and uptake in Africa: a systematic review and meta-analysis (1970-2024)": Addition Files 3

**Study period**

**Event rate (%)**

**Vaccine uptake (%)**

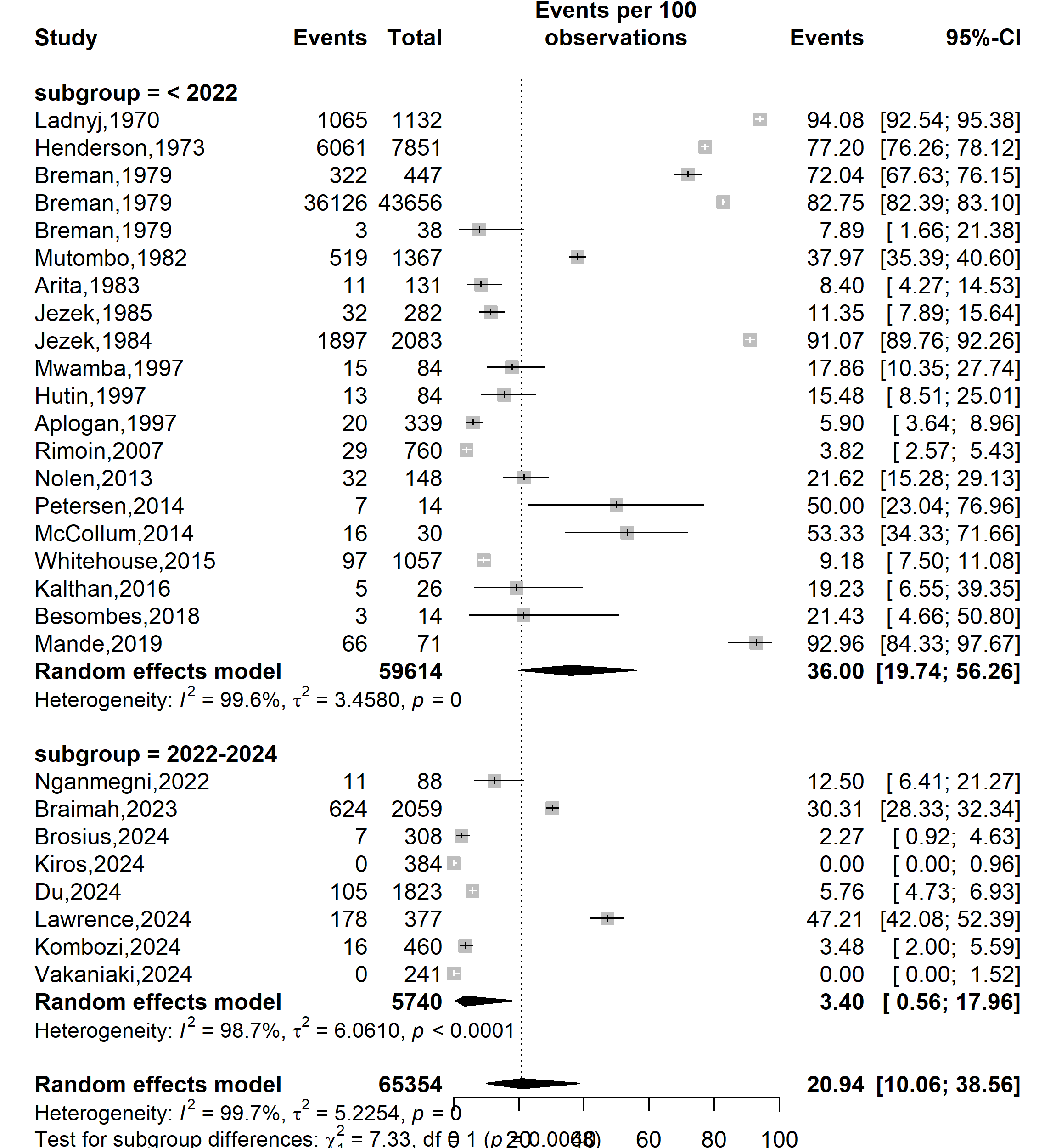

Supplementary Fig. 1 Pooled Mpox vaccine uptake rate by study periods in Africa, 1970-2024

**Country**

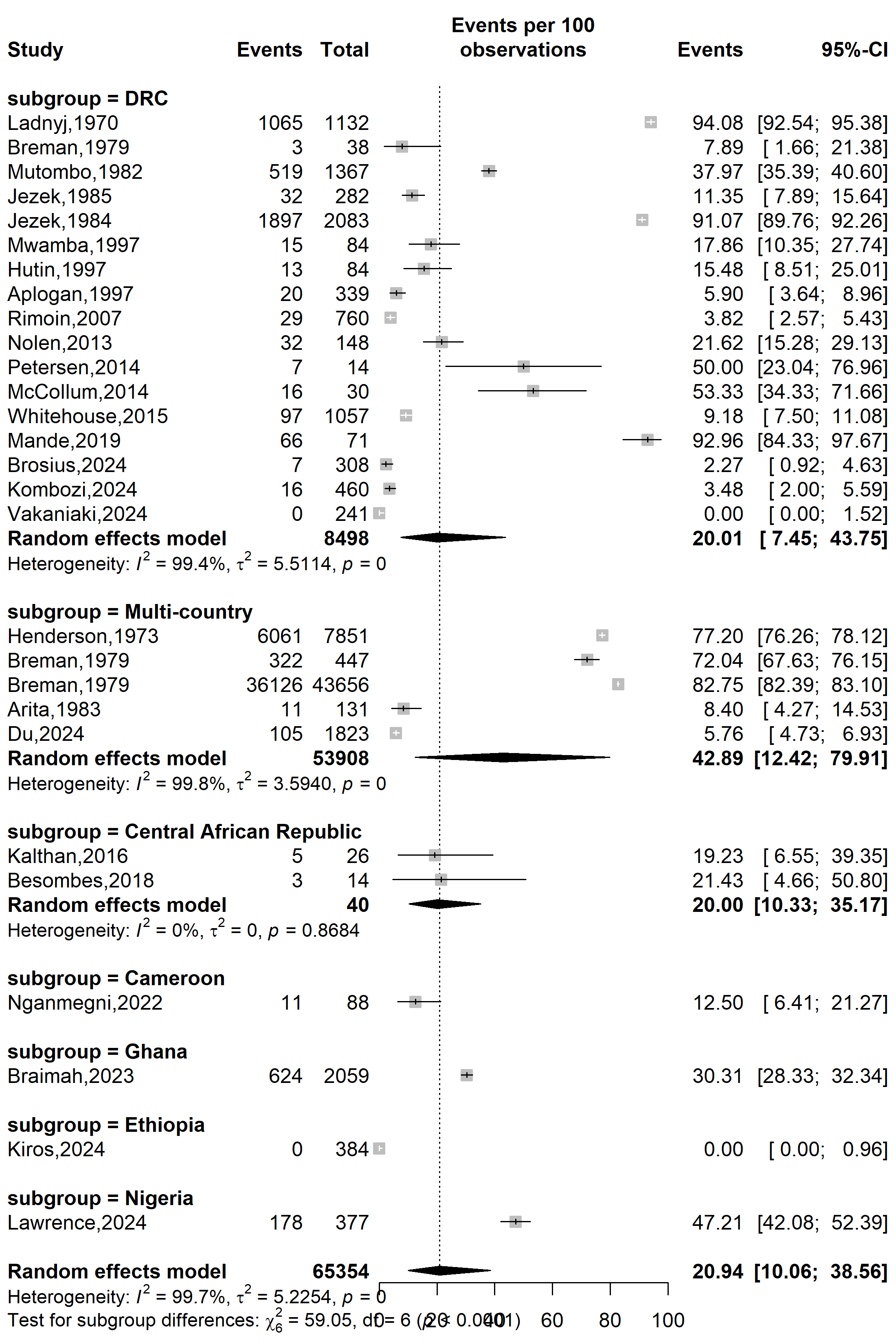

**Vaccine uptake (%)**

**Event rate (%)**

Supplementary Fig. 2 Pooled Mpox vaccine uptake rate by countries in Africa, 1970-2024

Abbreviation: DRC: Democratic Republic of Congo

**Type of participants**

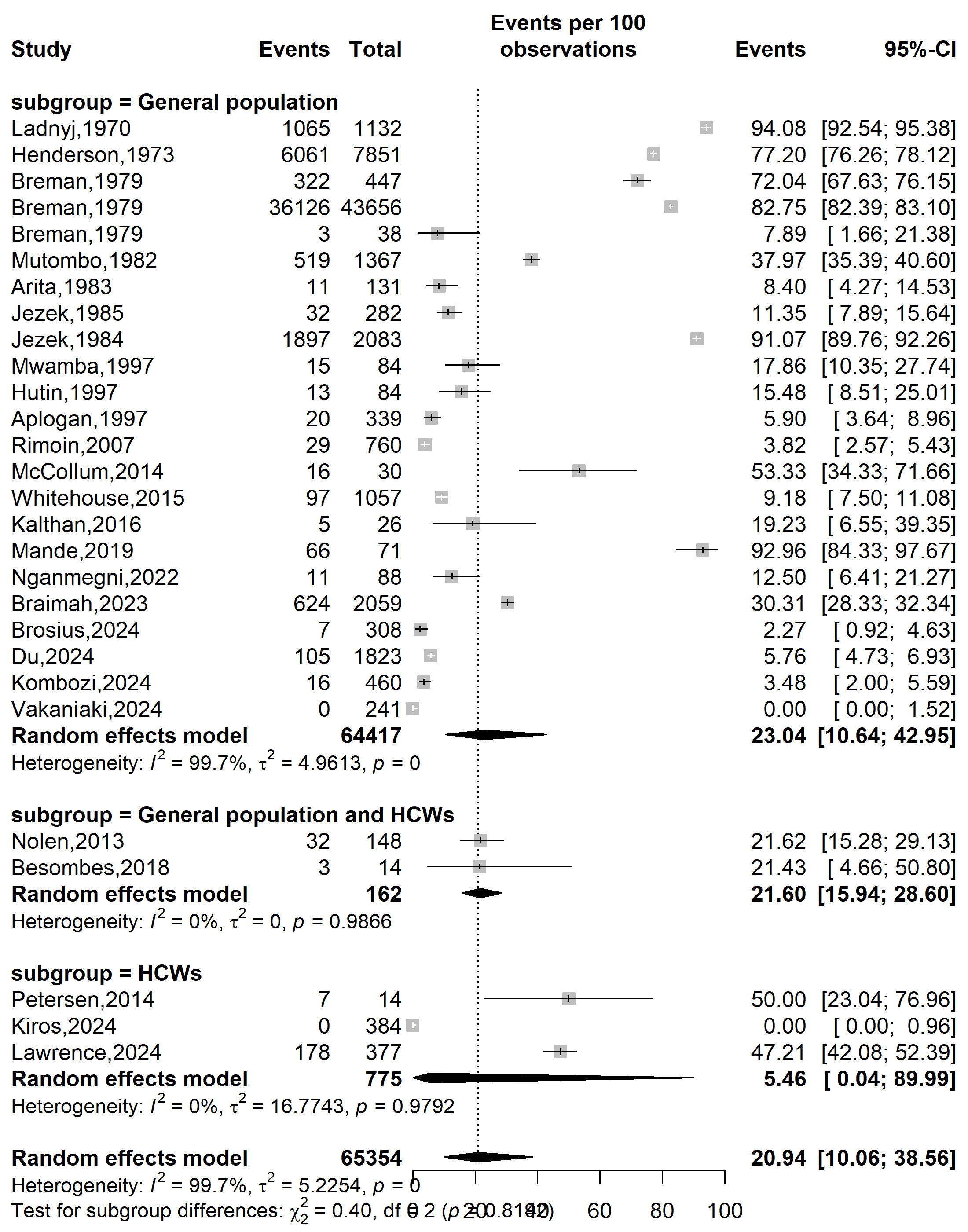

**Vaccine uptake (%)**

**Event rate (%)**

Supplementary Fig. 3 Pooled Mpox vaccine acceptance rate in Africa by types of participants, 2013-2024

Abbreviation: HCW: Healthcare worker

**WHO Afro zone**

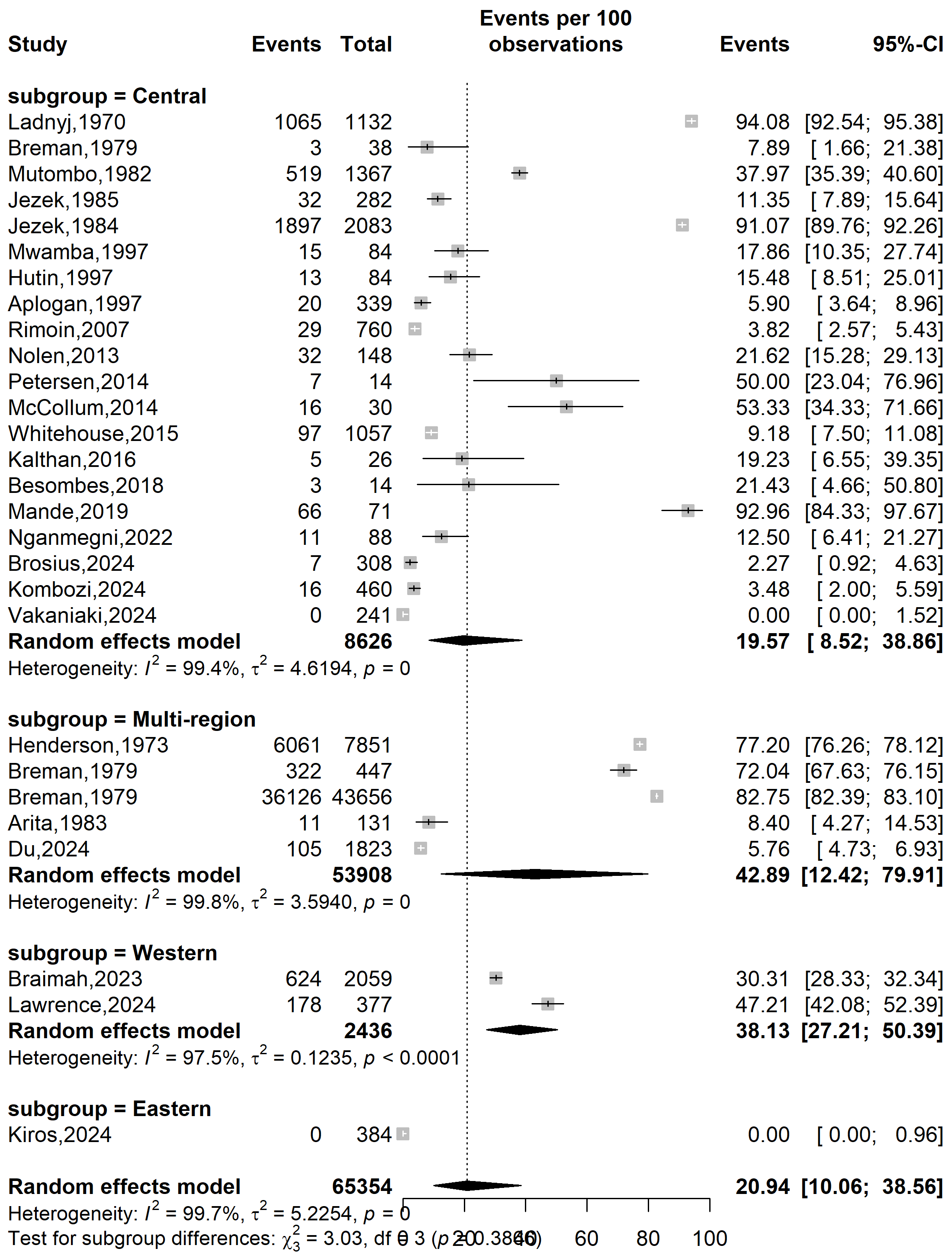

**Event rate (%)**

**Vaccine uptake (%)**

Supplementary Fig. 4 Pooled Mpox vaccine uptake rate in Africa by WHO’s country categorization, 1970-2024

**Setting**

**Event rate (%)**

**Vaccine uptake (%)**

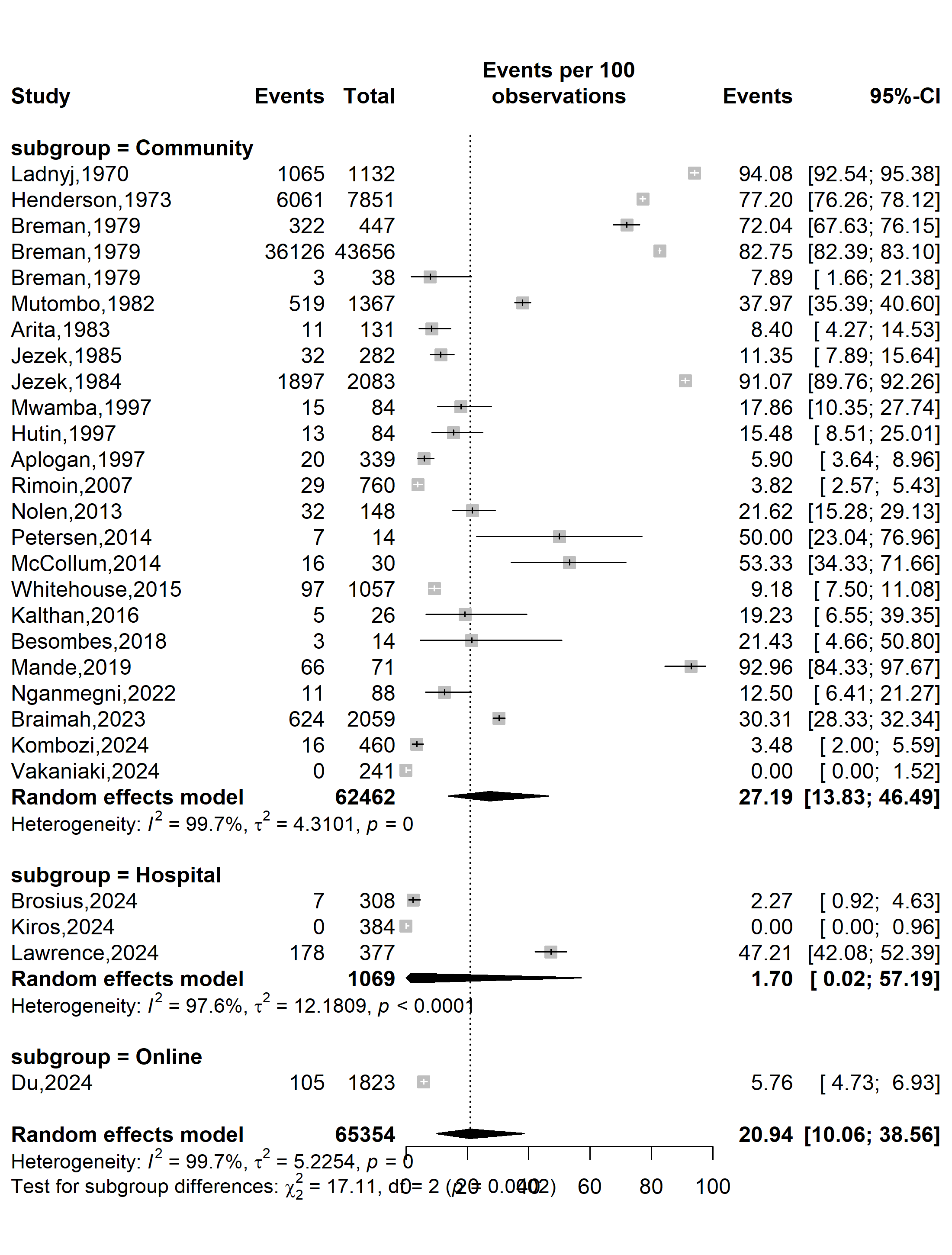

Supplementary Fig. 5 Pooled Mpox vaccine uptake rate in Africa by study setting, 1970-2024

**Type sampling**

**Vaccine uptake (%)**

**Event rate (%)**

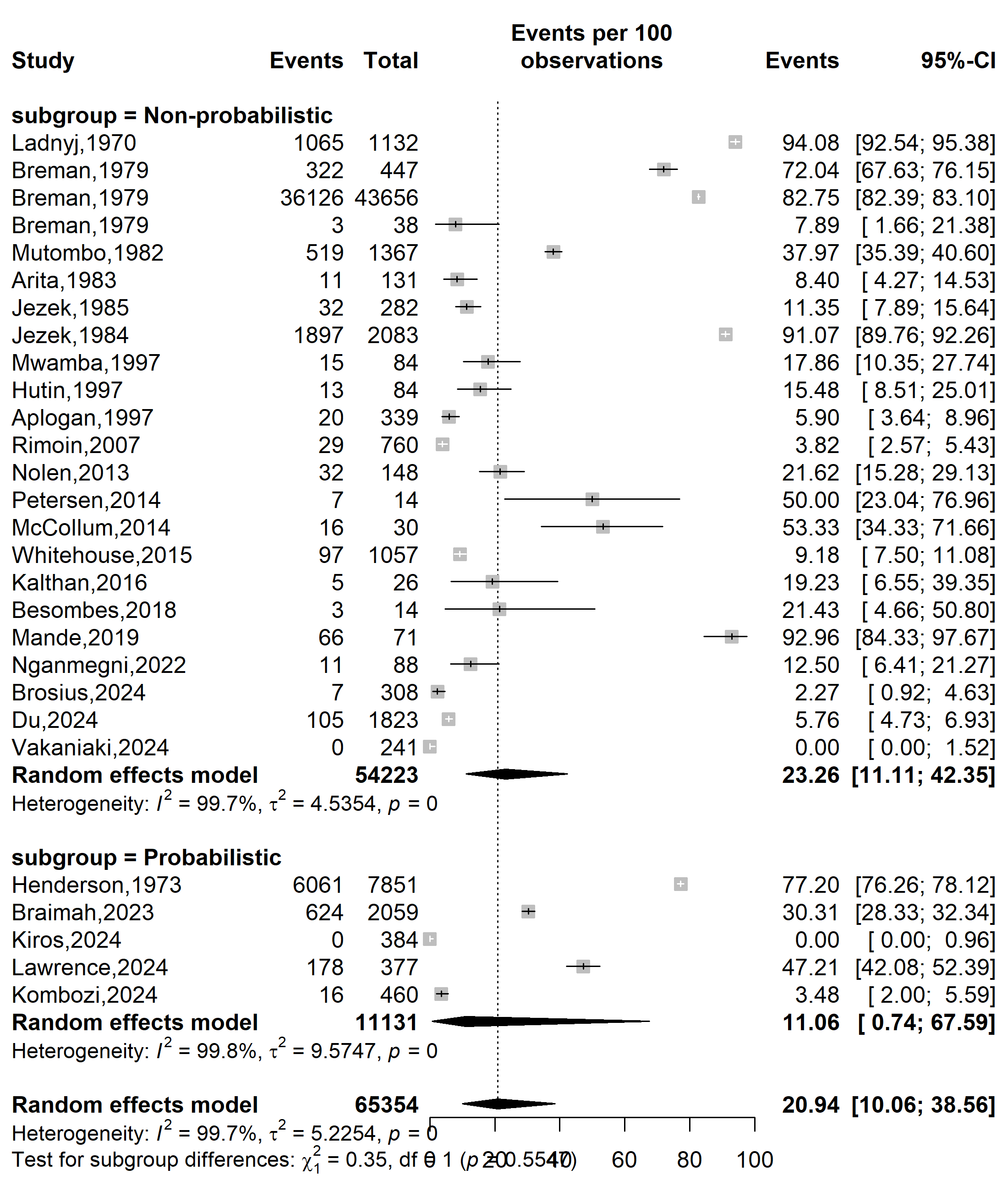

Supplementary Fig. 6 Pooled Mpox vaccine uptake rate in Africa by sampling method, 1970-2024

**Sample size**

**Vaccine uptake (%)**

**Event rate (%)**

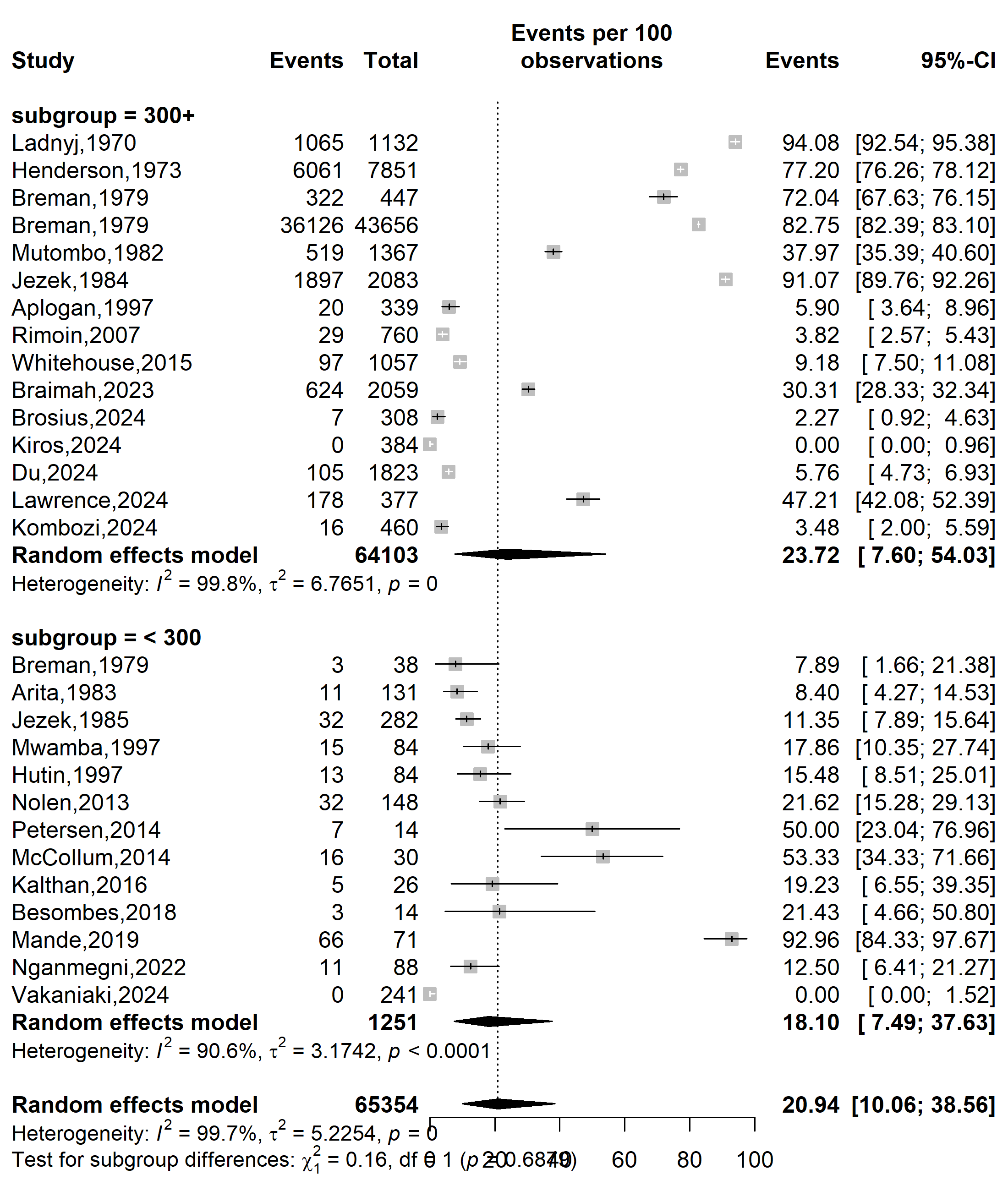

Supplementary Fig. 7 Pooled Mpox vaccine uptake rate in Africa by sample size, 1970-2024

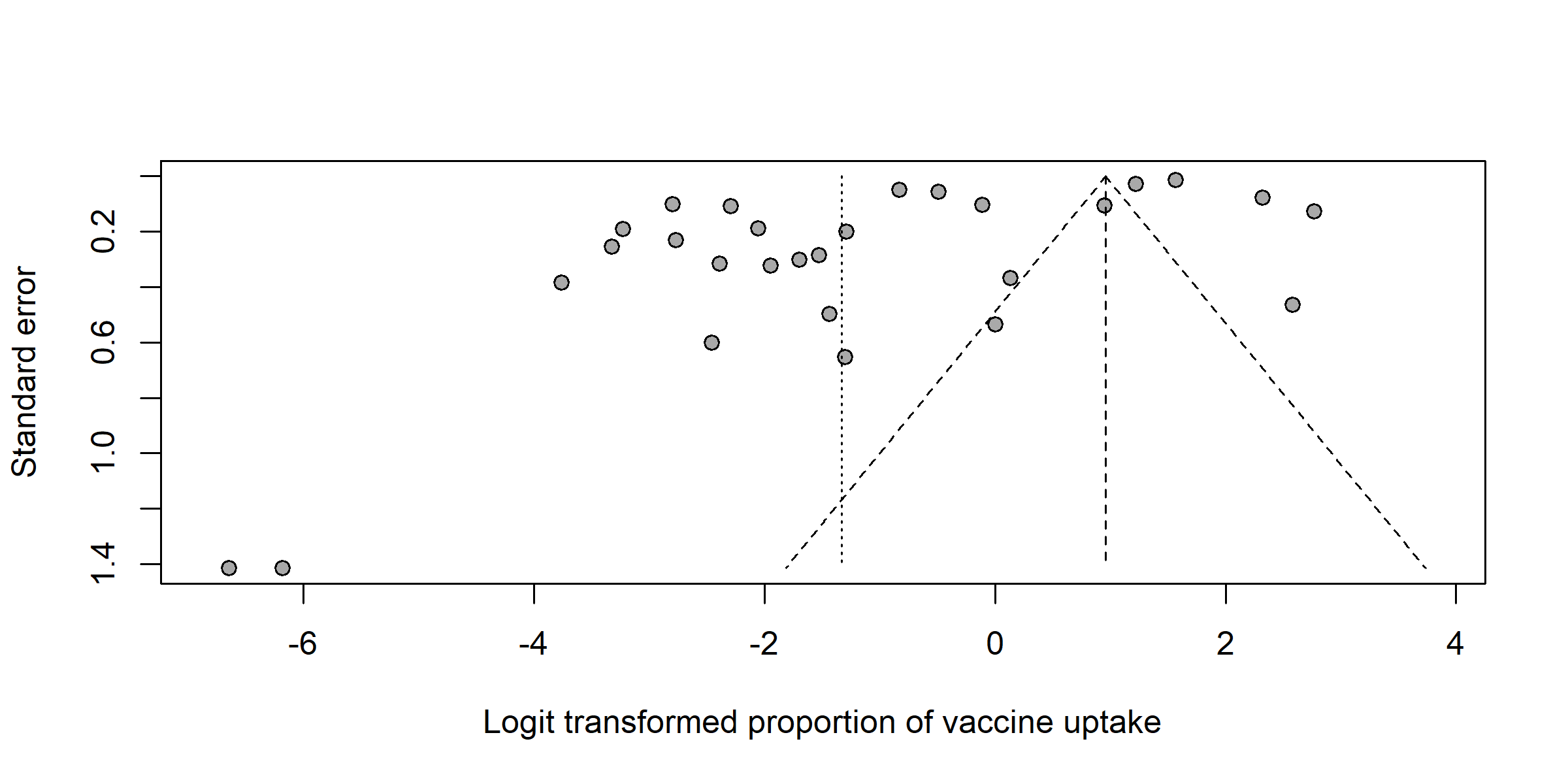

Egger’s test *p*-value < 0.001

Begg’s test *p*-value = 0.143

Supplementary Fig. 8 Funnel plot displaying the pseudo 95% confidence limits and tests assessing the publication bias of studies included, 1970-2024

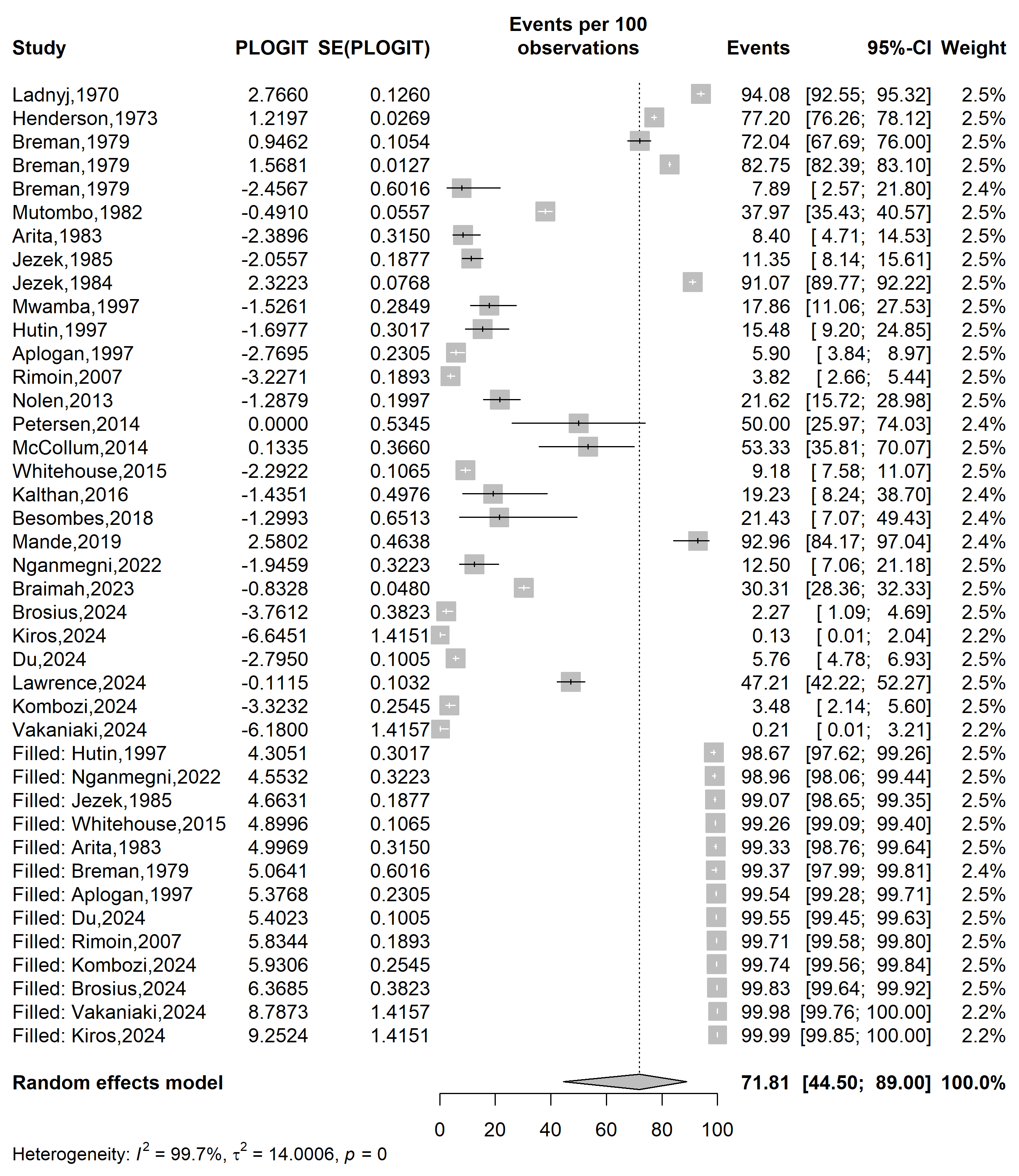

**Event rate (%)**

**Vaccine uptake (%)**

Supplementary Fig. 9 Trim and fill plot of the vaccine uptake rate in Africa, 1970-2024

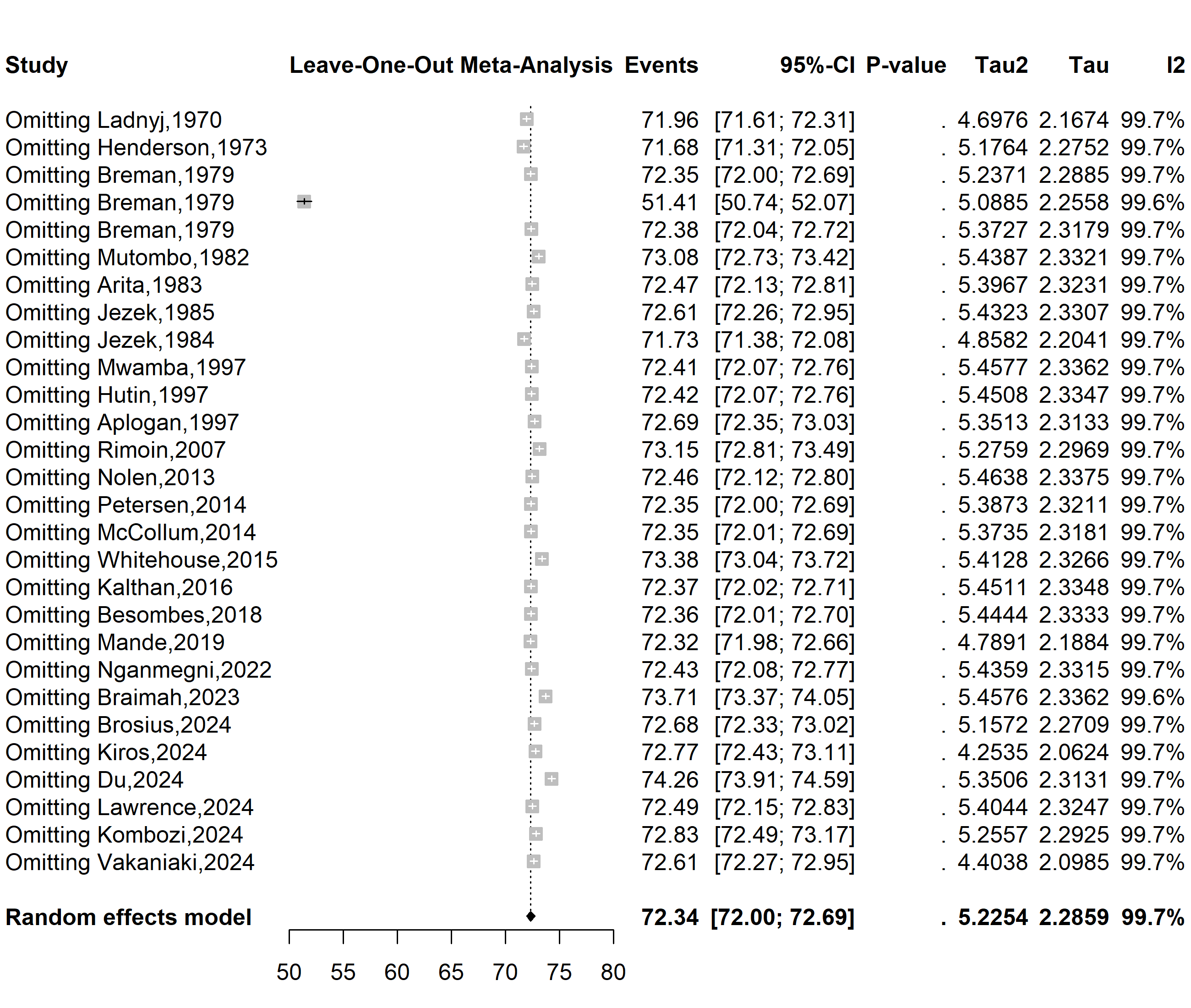

**Supplementary Fig. 10** Sensitivity analysis of the Mpox vaccine uptake in Africa, 2013-2024
